# Genetic drivers and clinical consequences of mosaic chromosomal alterations in 1 million individuals

**DOI:** 10.1101/2025.03.05.25323443

**Authors:** Kun Zhao, Yash Pershad, Hannah M Poisner, Xiaolong Ma, Kali Quade, Weizhi Cao, Liying Xue, Caitlyn Vlasschaert, Taralynn Mack, Nikhil K Khankari, Kelly von Beck, James Brogan, Ashwin Kishtagari, Robert W Corty, Yajing Li, Yaomin Xu, Bjoernar Tuftin, Laura Raffield, Stephen S Rich, Jerome I Rotter, Robert E Gerszten, Michael Y Mi, Alexander P Reiner, Paul Scheet, Paul L Auer, Alexander G Bick

## Abstract

Mosaic chromosomal alterations of the autosomes (aut-mCAs) are large structural somatic mutations which cause clonal hematopoiesis and increase cancer risk. Here, we detected aut-mCAs in 1,252,761 participants across four biobanks. Through integrative analysis of the minimum critical region and inherited genetic variation, we nominated candidate driver genes for each aut-mCA, prioritizing proto-oncogenes within recurrently gained regions and tumor suppressors within recurrently lost regions. We identified three novel inherited risk loci in *MAD1L1*, *TCL1A*, and *ATP2A3* that modulate aut-mCA risk and four novel aut-mCA-specific loci. We found specific aut-mCAs are associated with cardiovascular, cerebrovascular, or kidney disease incidence. High-risk aut-mCAs were associated with elevated plasma protein levels of therapeutically actionable targets: NPM1, PARP1, and TACI. Participants with multiple high-risk features such as high clonal fraction, more than one aut-mCA, and abnormal red cell morphology had a 50% cumulative incidence of blood count abnormalities over 2 years. Leveraging inherited variation, we provide evidence supporting a role for specific autosomal mCAs in chronic lymphocytic leukemia development. Together, our findings provide a framework integrating somatic mosaicism, germline genetics, and clinical phenotypes to identify individuals who could benefit from preventative interventions.

## Introduction

As individuals age, hematopoietic stem cells (HSCs) accumulate somatic mutations, some of which enhance HSC fitness and result in peripheral blood cells sharing the same mutation. This phenomenon, termed clonal hematopoiesis, affects 10% of people over the age of 60 and increases risk of all-cause mortality, hematologic malignancy, cardiovascular disease, and infection.^1–7^ One way to classify clonal hematopoiesis is by the type of driver mutation detectable from blood-derived DNA. Single-nucleotide variants and small insertions or deletions in certain preleukemic genes are termed clonal hematopoiesis of indeterminate potential (CHIP).^1,8–10^ Larger >1 megabase gains (+), losses (-), or copy-neutral losses of heterozygosity (=) are termed mosaic chromosomal alterations (mCAs).^11–16^ mCAs of sex chromosomes such as mosaic loss of Y and loss of X are the most common type of mCA and have been extensively studied.^17,18^ In contrast, mCAs of the autosomes (aut-mCAs) are more heterogeneous in type and phenotypic consequence.^4,19–21^

Prior work has shown that aut-mCAs increase in prevalence with age, the presence of aut-mCAs is influenced by inherited genetic variants, and aut-mCAs often co-occur with blood count abnormalities and increase risk of chronic lymphocytic leukemia (CLL) and severe infections.^3,14,19,22^ However, several important questions about aut-mCAs remain unanswered. First, for most aut-mCAs, the genetic determinants underlying their selective advantage remain incompletely understood, and may involve the combined effects of multiple genes across the affected chromosomal region.^15^ Second, although recent studies have begun to examine mCAs in ancestrally diverse populations, including African, Hispanic, and East Asian cohorts, most analyses have remained limited in sample size or focused on specific ancestries.^11,19,23^ This has constrained comprehensive evaluation of how germline genetics influence aut-mCA prevalence across populations. Third, unlike in CHIP where it is well- known that different driver genes have distinct risk factors and phenotypic consequences, researchers have not had sufficient sample size to profile the heterogeneity of specific aut-mCAs. Fourth, while certain aut-mCAs are associated cross-sectionally with abnormal blood counts,^11,14,15^ for a person with an aut-mCA but normal blood counts, it is not well-understood what their risk of developing incident blood count abnormalities, the earliest sign of progression to hematologic malignancy. Fifth, no established preventive therapies are available to reduce the risk of progression.

Here, using 1,252,761 individuals across four large-scale biobanks, NIH All of Us (AoU) version 8, NHLBI TOPMed, Vanderbilt’s BioVU, and UK Biobank (UKB) with genetic and longitudinal clinical and laboratory data, we have constructed the largest and most diverse, deeply phenotyped cohort of aut-mCAs (**Extended Data Fig. 1**). Using this data, we identified candidate drivers for all aut-mCAs, discovered novel aut-mCA-specific and aut-mCA-wide germline genetic variants which alter risk of aut-mCAs, and reported the proteomic signature of aut-mCAs to nominate candidate biomarkers and therapeutic targets. We are the first to delineate the health consequences of specific aut-mCA types beyond hematologic malignancy. Then, we identified a subgroup of people with aut-mCAs at high risk of incident blood count abnormalities. Finally, we provide evidence that aut-mCAs are associated with increased risk of CLL and that polygenic germline risk and somatic aut-mCAs interact to alter CLL risk.

## Results

### The profile of autosomal mosaic chromosomal alterations

We profiled aut-mCAs in 1,252,761 individuals using genotyping arrays in UKB^14,15^ and AoU^24^ and whole genome sequencing in BioVU^25^ and TOPMed^11^ (**Extended Data Fig. 2**). The baseline characteristics and mCA detection of each cohort are shown in **Supplementary Table 1-2.** The participants in the study had a mean age at DNA collection of 53 years; the mean age of those with an aut-mCA was 62 years. 42% of participants were male. 77% clustered with the 1000 Genomes European ancestry group (1KG-EUR-like), 12% with the 1000 Genomes African ancestry group (1KG-AFR-like), and 2% with the 1000 Genomes East Asian ancestry group (1KG-EAS-like). 30,782 (2.5%) individuals had ≥1 detectable aut-mCA. Among those aut-mCAs, 70% had one, 13% had two, 17% had multiple (≥3) (**Extended Data Fig. 3**). The prevalence of aut-mCA increased with age, with 1KG-EUR-like and 1KG-AFR-like populations exhibiting higher detection rates of aut-mCAs than 1KG- EAS-like populations in each age group (**Extended Data** Fig.4). We note that reduced haplotype phasing accuracy in EAS populations may influence mCA detection. However, results were consistent in a sensitivity analysis restricted to higher cell fraction mCAs (>5%), for which phasing-related bias is less likely to affect detection (**Extended Data Fig. 5**).

Certain mCA subtypes were markedly overrepresented in different age and sex groups. A majority of people with chr15+ (70%) aut-mCAs were males and 79% of those with chr10q- were female (**Figure 1A**). The sex bias of chr15+ and chr10q– persists after exclusion of those with mosaic loss of chromosome X and Y (**Extended Data Fig. 6)**. Although this finding of chr15+ had previously been reported by UKB and TOPMed,^11,15^ we further validated this observation using data from AoU and BioVU. People with CN-LOH aut-mCAs were younger on average than those with gain (*P* = 7×10^-94^) or loss aut-mCAs (*P* = 7×10^-30^). The prevalence of certain autosomal mCAs varied by genetic ancestry. For example, chr15+ were more common in individuals of 1KG-EUR-like ancestry than in those of 1KG-AFR-like ancestry (3.2% vs. 1.3%; *P* = 5.0×10^-18^), whereas chr11q– events were more frequent in 1KG-AFR-like individuals (3.0% vs. 1.4%; *P* = 9.4×10^-13^) (**Figure 1B, Extended Data Fig. 7**).

**Figure 1.**
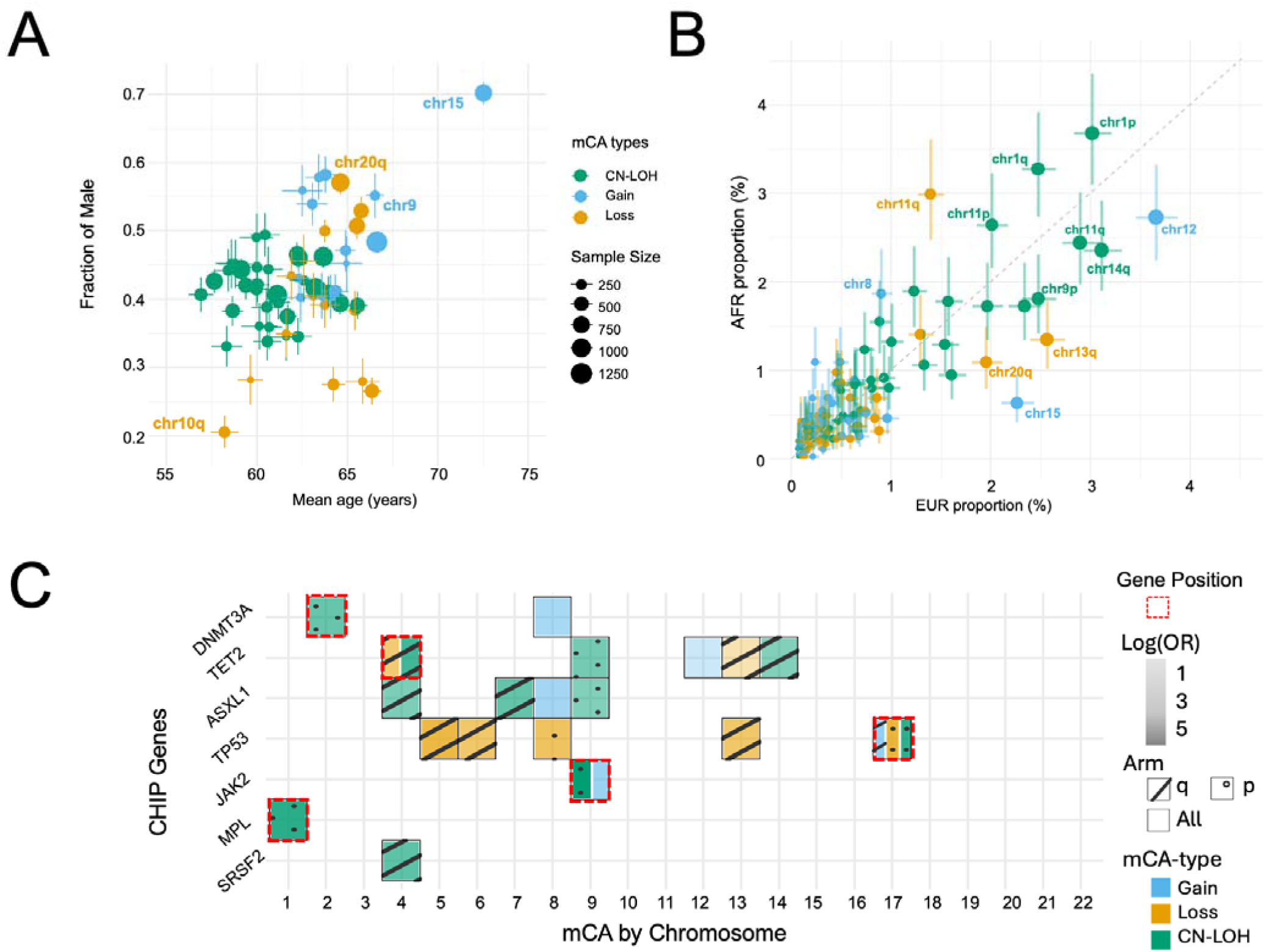
Demographic and genetic patterns of autosomal mosaic chromosomal alterations. A) Age and sex distribution of aut-mCA types from 30,782 unrelated mCA cases. Each point represents a specific aut-mCA type, with x-axis showing mean age of cases and y-axis showing proportion of males. B) Ancestry-specific aut-mCA frequencies comparing 1KG-EUR-like and 1KG-AFR-like populations. C) Co-occurrence patterns between CHIP mutations and aut-mCAs across chromosomes. Color indicates aut- mCA type (CN-LOH, gain, loss). Firth logistic regression was performed and the model was adjusted for age, sex, and cohort to ensure robust results. Furthermore, co-occurrence pairs with fewer than 10 individuals were excluded, and a Bonferroni correction (*P* < 0.05/180) was applied to account for multiple testing.

### CHIP and autosomal mosaic chromosomal alterations co-occurrence

Given that co-occurring somatic mutations may influence the prevalence of aut-mCAs, we characterized the patterns of co-occurrence between CHIP mutations and aut-mCAs by Firth logistic regression. Among the aut- mCA carriers, 2,692 (8.7%) also had detectable CHIP. CHIP mutations tended to co-occur with = aut-mCAs of the same chromosome (**Figure 1C**), such as *TET2*-chr4= (OR: 20.6, 95% CI 17.0-24.9, *P* < 1×10^-300^), *MPL*- chr1= (OR: 53.7, 95% CI 27.4-109.0, *P* < 1×10^-300^), *DNMT3A*-chr2= (OR: 5.7, 95% CI 4.3-7.6, *P* = 1×10^-31^), and *TP53*-chr17= (OR: 42.6, 95% CI 27.9-64.9, *P* < 1×10^-300^) (**Supplementary Table 3, 4**). In addition, *TP53* was associated with multiple aut-mCA losses, while chr8+ was associated with *DNMT3A*-CHIP (*P* = 3×10^-5^). The results remained robust in a sensitivity analysis limited to co-occurring CHIP and mCA events where both had cell fractions greater than 5% (**Extended Data Fig. 8)**.

### Prioritization of candidate driver genes

For most aut-mCAs, the driver gene conferring their selective advantage remains unknown. To investigate whether aut-mCAs are shaped by selection acting on cancer driver genes, we first quantified the enrichment of known hematologic malignancy genes within aut-mCA regions. Consistent with models of cumulative haploinsufficiency and triplosensitivity observed in cancer aneuploidy,^26,27^ we found that regions of copy number loss were significantly enriched for tumor suppressor genes (permutation *P* = 0.001) and regions of gain were enriched for proto-oncogenes (permutation *P* = 0.003).

Building on this observation, we prioritized specific candidate drivers by identifying the minimum shared altered region (MSAR) for each aut-mCA type across all cohorts (Methods; **Supplementary Table 5**). Within these regions, we prioritized genes implicated in hematologic malignancy that were biologically consistent with the copy number change. Applying this prioritization, we identified candidate proto-oncogene drivers within recurrently gained regions and candidate tumor-suppressor drivers within recurrently lost regions; CN-LOH regions were not restricted by gene class and yielded candidates of both classes (**Table 1**). For example, the MSAR of chr11 alterations (gain, loss, and CN-LOH) contained *WT1*, which may act as both a tumor suppressor and proto-oncogene in a context-specific manner.^28^ Many aut-mCA putative drivers were genes implicated in CHIP such as *DNMT3A*, *TET2*, *JAK2*, *ASXL1*, *SF3B1*, *TP53*, and *RUNX1*.

**Table 1.**
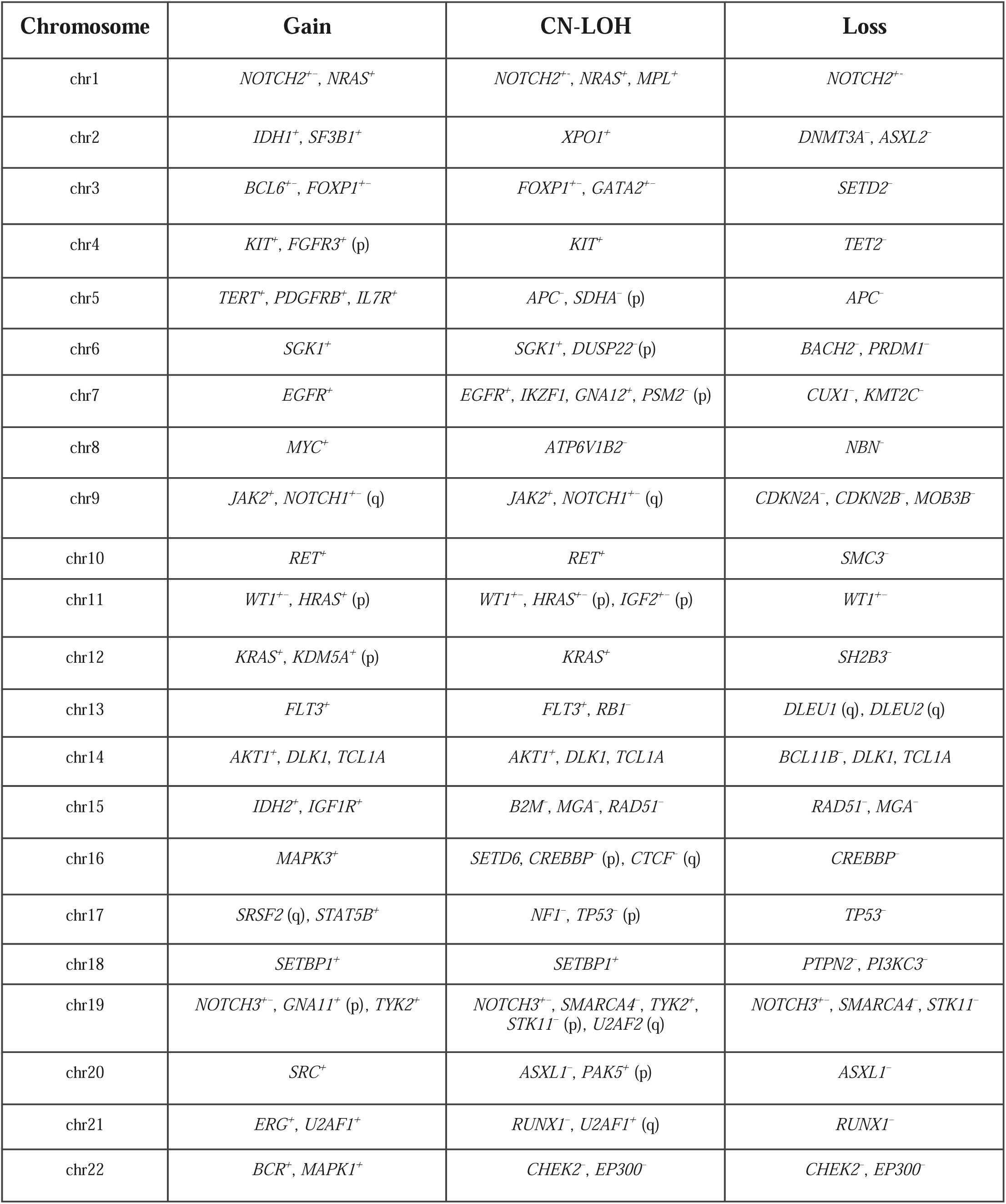
Candidate driver genes associated with autosomal mosaic chromosomal alterations. Genes are annotated as proto-oncogenes (+), tumor suppressors (-), or both (+/-). Genes were identified from the minimum shared altered region in each chromosomal event type and filtered for those implicated in hematologic malignancy. Annotations in parentheses (p) and (q) indicate chromosome arm specificity when relevant. CN-LOH = copy-neutral loss of heterozygosity. Genes are italicized.

Our genomic region analysis also highlighted unique features of chr10q–. While this event has been previously linked to germline fragility at *FRA10B*,^15^ this site is not known to drive malignancy and is unlikely to confer clonal fitness. However, the deletion encompasses the tumor suppressor *SMC3*. Loss of *SMC3* results in profound genomic instability, which may explain the unique fitness landscape of 10q– clones and their potential limitations in transforming to malignancy.^21,29^

### Chromosome-arm-localized germline genetic determinants of autosomal mCAs

We hypothesized that common germline variants, distinct from rare driver mutations, significantly alter the risk of specific aut-mCAs and may facilitate clonal selection. In a meta-analysis of the 4 cohorts after ancestry- stratified association testing, we identified 10 significant *cis* associations (i.e., genetic variants associated with the specific aut-mCA of the chromosome and arm where the variant resides) after multiple hypothesis correction (**Supplemental Table 6**). These associations spanned all alteration types: copy number losses (–), copy-neutral loss of heterozygosity (=), and copy number gains (+). To rule out genotyping artifacts and confirm that these variants drive clonal selection, we analyzed phased genotyping data from AoU version 8 and TOPMed. We observed significant allelic shift (P<5×10^-8^) for all *cis*-variants, confirming that the chromosome carrying the risk allele is preferentially amplified or retained in the clonal population (**Supplemental Table 6**). Six known associations with variants near *MPL* (chr1p=, β = 1.01, standard error (SE) = 0.14, *P* = 2×10^-13^), *JAK2* (chr9p=, β = 0.85, SE = 0.06, *P* = 3×10^-48^), *ATM* (chr11q=, β = 0.28, SE = 0.04, *P* = 5×10^-12^), *TM2D3* (chr15q=, β = 0.74, SE = 0.08, *P* = 2×10^-20^), *DLK1* (chr14q=, β = 0.41, SE = 0.05, *P* = 4×10^-16^), and *FRA10B* (chr10q–, β = 2.13, SE = 0.10, *P* = 2×10^-93^) were significant in our meta-analyses.

Our meta-analysis revealed four novel variant associations. All four implicated genes were expressed across hematopoietic stem and progenitor populations, including hematopoietic stem cells, multipotent progenitors, and common lymphoid progenitors (**Extended Data Figure 9)**. To ensure the validity of our hits, we tested the association between the significant loci and the same type of mCA of the opposite arm and found no evidence of a significant association. The inclusion of more participants of 1KG-AFR-like ancestry enabled discovery of many variants with different minor allele frequencies by ancestry. A missense variant in *CLN5* causing amino acid change K319R was associated with increased risk of 13q=, a high-risk CLL-associated mCA (β = 0.34, SE = 0.06, *P* = 5×10^-10^); the variant is much more common in 1KG-AFR-like than 1KG-EUR-like individuals (minor allele frequency of 0.47 versus 0.10) and was thus likely underpowered in prior analyses in the UK Biobank. Similarly, variants near *TMEM70* (chr8q+, β = 0.35, SE = 0.05, *P* = 8×10^-12^), *PIK3C3* (chr18q=, β = 2.48, SE = 0.40, *P* = 6×10^-10^), and *FBXO39* (chr17p–, β = 0.93, SE = 0.14, *P* = 3×10^-11^) are more common in 1KG-AFR-like than 1KG-EUR-like individuals. *TMEM70* encodes a mitochondrial membrane protein essential for ATP synthase assembly which supports the metabolic demands of cellular differentiation; the association of rs73687116-C with chr8q+ acquisition suggests that the variant may provide a proliferative advantage to lymphoid progenitors with chr8q+. *FBXO39* encodes a FBOX-family ubiquitin ligase; this family of proteins has been implicated in leukemia due to their role in regulation of the cell cycle in hematopoiesis. *PIK3C3* encodes a class III phosphatidylinositol 3-kinase essential for autophagy initiation and lineage commitment of hematopoietic stem and progenitor cells.

### Germline genetic risk of developing any autosomal mosaic chromosomal alteration

In contrast to germline variants on the same chromosome altering risk of specific aut-mCAs, certain germline genetic variants altered the risk of any detectable aut-mCA. We identified five independently-associated loci, three of which were novel (**Figure 2A**). Two loci which had been previously shown to be associated with aut- mCA risk in the UK Biobank by Loh et al^14^ are (1) rs7705526-C (OR: 0.90, 95% CI 0.88-0.92, *P* = 1×10^-20^, 1KG-EUR-like allele frequency (AF) = 0.33, 1KG-AFR-like AF = 0.19) on chr5 in *TERT* and (2) rs1356532206-TA (OR: 1.1, 95% CI 1.05-1.10, *P* = 8×10^-9^, 1KG-EUR-like AF = 0.42, 1KG-AFR-like AF = 0.67) on chr2 in *SP140* (**Supplementary Table 7**). The other three novel loci associated with increased aut- mCA prevalence were: (1) rs7792631-T (OR: 0.95, 95% CI 0.93-0.97, *P* = 4×10^-9^, 1KG-EUR-like AF = 0.30, 1KG-AFR-like AF = 0.24) in *MAD1L1* on chr7, (2) rs2887399-T (OR: 1.06, 95% CI 1.04-1.09, *P* = 1×10^-9^, 1KG-EUR-like AF = 0.21, 1KG-AFR-like AF = 0.40) on chr14 in *TCL1A;* and (3) rs8074479-C (OR: 0.91, 95%CI 0.88-0.95, *P* = 4×10^-9^, 1KG-EUR-like AF = 0.20, 1KG-AFR-like AF = 0.46) in *ATP2A3* on chr17. These significant loci did not exhibit a strong association with any particular aut-mCA, but rather were moderately associated with increased risk of multiple aut-mCAs (**Figure 2B**).

**Figure 2.**
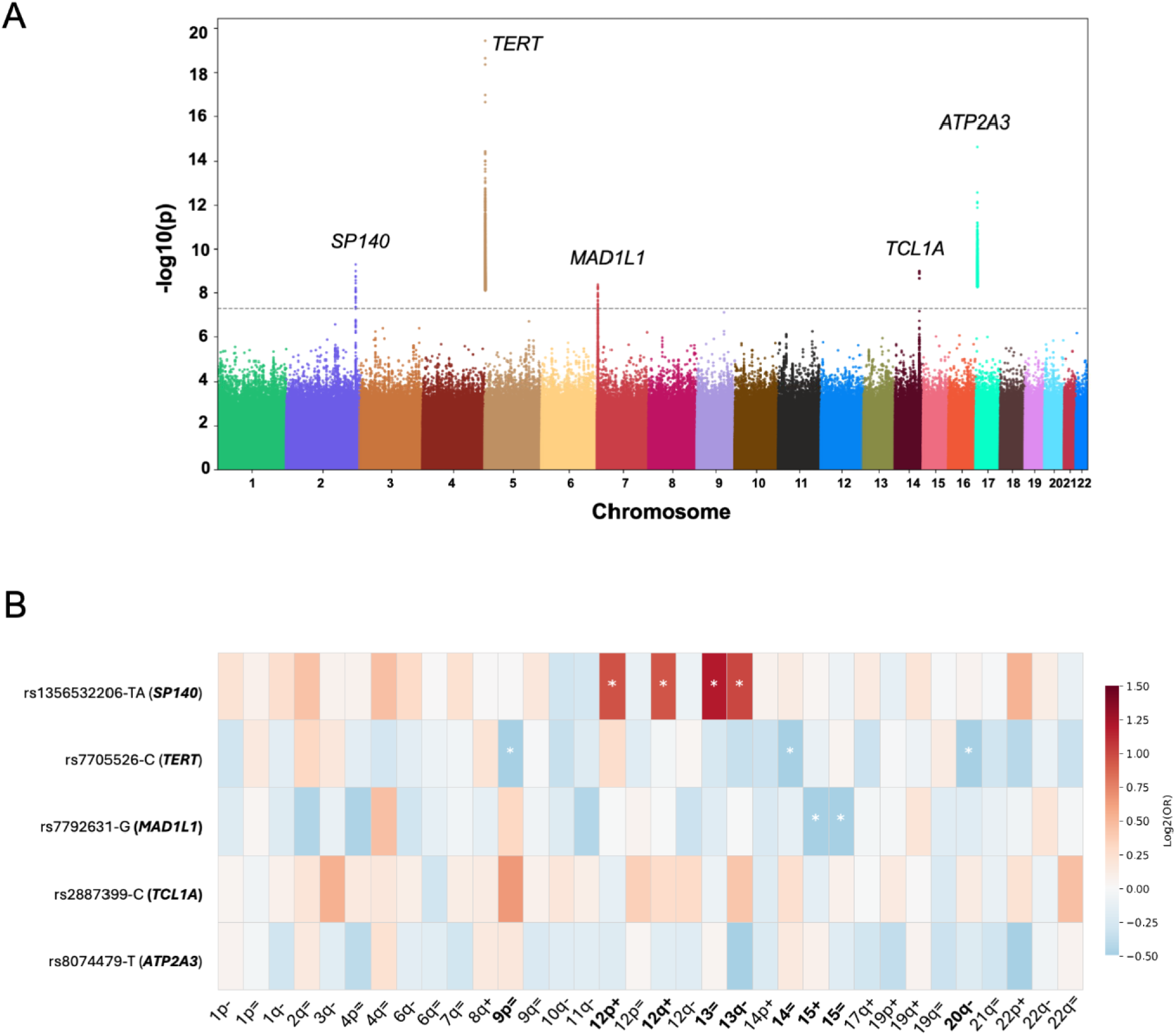
Genome-wide association analysis of autosomal mCA risk. A) Manhattan plot showing genome-wide significant loci (P<5×10^-8^) associated with aut-mCA prevalence. B) Heatmap showing the associations between five significant germline genetic loci identified in trans-GWAS by specific types of aut-mCAs. The rows represent the SNPs along with their corresponding genes, while the columns indicate different mCA types. The color scale represents the effect estimates, with red indicating positive associations and blue indicating negative associations. Asterisks (*) denote statistically significant associations at a nominal P value threshold of 0.05.

Given that aut-mCAs are enriched for CLL-associated lymphoid events and that our aggregate risk loci implicate lymphoid-restricted genes such as *SP140* and *TCL1A*, we assessed which blood cell types or states underlie the inherited risk of acquiring any aut-mCA. We employed the SCAVENGE method, which overlaps fine-mapping posterior probabilities derived from GWASs of aut-mCA susceptibility with accessible chromatin enrichments at single-cell resolution by employing a network propagation approach, using the Corces et al.^30^ reference of sorted hematopoietic populations. In contrast to mLOY and other myeloid-biased clonal hematopoiesis phenotypes^18^, in which enrichment localizes to hematopoietic stem cells and early progenitors, aut-mCA risk variants were most strongly enriched in mature B cells (mean trait-relevance score [TRS] 4.02; 89.5% of cells significantly enriched) (**Extended Data Figure 10**). A substantial component of inherited aut- mCA risk acts within the mature B-cell compartment rather than exclusively at the level of the stem cell, a notable divergence from the HSC-restricted architecture reported for mLOY and other forms of clonal hematopoiesis.

### Genetically predicted leukocyte telomere length and autosomal mCAs

Given that variants in *TERT* alter aut-mCA and CLL risk, we hypothesized that genetically predicted leukocyte telomere length (gLTL) may alter risk of aut-mCAs (**Supplementary Table 8**). The prevalence of aut-mCAs varied by *TERT* genotypes and gLTL stratification across age groups (**Extended Data Fig. 11**). Across all age groups, individuals with the T/T genotype exhibited the highest prevalence, whereas those with the C/C genotype had the lowest prevalence. Individuals with the highest gLTL (top 20%) had the highest aut-mCA prevalence. The difference in aut-mCA prevalence between gLTL groups was more pronounced among aut- mCAs considered high-risk for CLL^19,22^ (**Extended Data Fig. 12, Supplementary Table 9, 10**).

We further investigated the associations between specific mCAs and gLTL, and identified five specific types of aut-mCAs that were associated with longer gLTL, such as chr13q- (β = 1.23 , 95% CI 1.14-1.32, *P* = 9×10^-8^), chr9p= (β = 1.22, 95% CI 1.13-1.32, *P* = 1×10^-6^), and chr11q- (β = 1.25, 95% CI 1.12-1.38, *P* = 4×10^-5^) after correction for multiple hypothesis testing **(Supplementary Table 11)**. In addition, we analyzed the association between measured LTL and aut-mCAs in UK Biobank. We observed that shorter measured LTL was significantly associated with higher prevalence of overall aut-mCAs after adjustment (OR = 0.92, 95% CI: 0.90- 0.94, *P* = 5 ×10^-15^). In addition, we tested the association between specific aut-mCA and measured LTL, and found that five types were significantly associated with shorter LTL, including chr1q+, chr4q=, chr9+, chr9p=, and chr11q= **(Supplementary Table 11).**

### Proteomic associations with autosomal mCAs

In order to nominate potentially actionable therapeutic targets, we sought to identify the proteomic signature of aut-mCAs. Using the UK Biobank plasma proteomics data for 52,705 participants, we examined the associations between ∼1500 proteins and the presence of an aut-mCA. We identified several proteins with significant associations that overlap with targets of FDA-approved drugs, such as PARP1, NPM1, and TNFRSF13B (TACI) (**Supplementary Table 12**). Aut-mCAs (β = 0.25, SE = 0.01, *P* = 2×10^-73^) and high-risk CLL-associated aut-mCAs (β = 1.37, SE = 0.04, *P* = 9×10^-307^) were associated with increased Fc receptor-like 2 (FCRL2) levels (**Figure 3A**; **Figure 3B**). Several proteins from the tumor necrosis factor receptor superfamily (TNFRSF) also showed strong associations with aut-mCAs and high-risk CLL-associated aut-mCAs, including TNFRSF9 (β = 0.20-0.98, *P* < 1×10^-54^), TNFRSF13B (β = 0.11-0.47, *P* < 6×10^-29^), and TNFRSF13C (β = 0.13-0.83, *P* < 1×10^-26^). We validated our findings using OLINK proteomic data from the TOPMed MESA cohort (184 cases and 3,966 controls). Among the 73 proteins which are associated with aut-mCAs in UKB, 72 showed consistent directionality in MESA (sign test *P* = 1.57 × 10^-20^), and 21 of these reached nominal significance (**Supplementary Table 13**). Higher levels of PARP1, TCL1A, and NPM1 were also associated with aut-mCAs, lymphoid aut-mCAs, and high-risk CLL-associated aut-mCAs, with a stronger association for high-risk CLL- associated aut-mCAs. We then tested the association between proteins and cell fraction of individuals with mCAs, indicating the clonal expansion (**Supplementary Table 14**). FCER2, which is considered a positive prognostic marker for CLL^31^, exhibited the strongest association with the cell fraction of aut-mCAs (β = 2.34, SE = 0.17, *P* = 3×10^-40^) and lymphoid mCAs (β = 4.62, SE = 0.36, *P* = 8×10^-32^). Similarly, PARP1, TCL1A, and NPM1 were also associated with clonal fraction of aut-mCAs, lymphoid aut-mCAs, and high-risk CLL- associated aut-mCAs. Proteins associated with presence of high-risk CLL-associated aut-mCAs were enriched for GO biological processes involving activation of T cells (N = 8 proteins, *P* = 6.4×10^-7^) and B cells (N = 7 proteins, *P* = 8.5×10^-7^).

**Figure 3.**
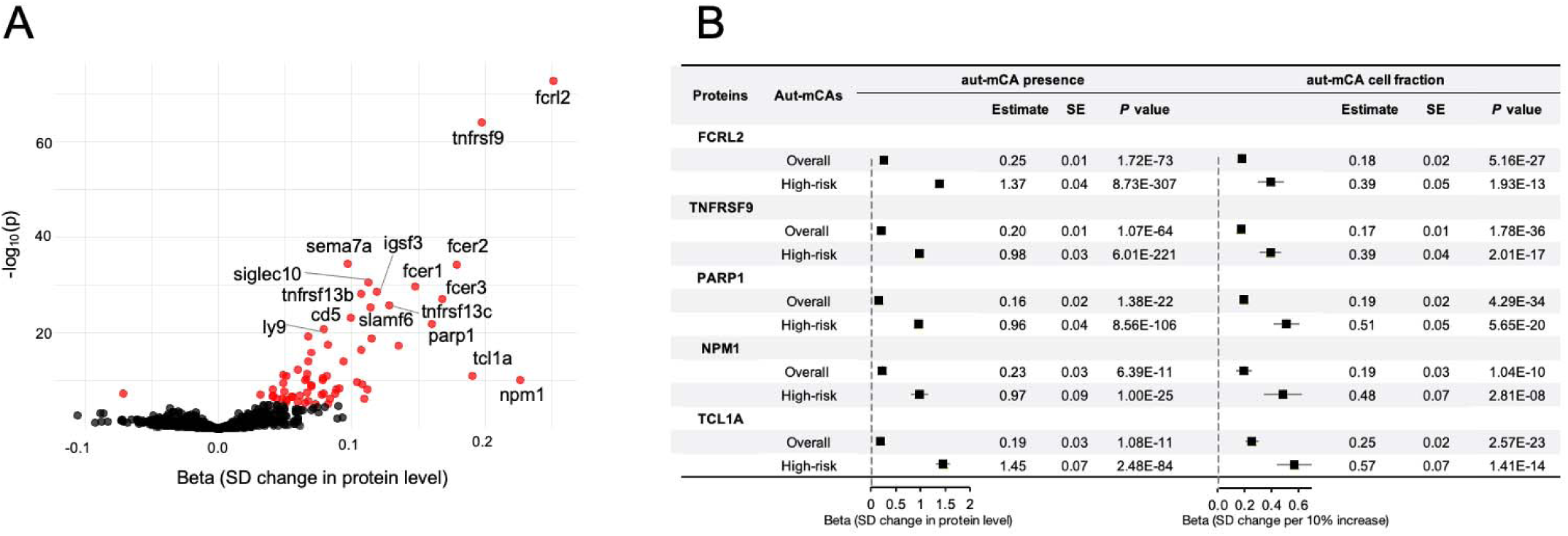
Proteomic signatures of autosomal mCAs. (A) Volcano plot showing associations between protein levels and the presence of autosomal mCAs. The x-axis represents the regression coefficient (beta), corresponding to the change in standardized protein levels (SD units) associated with autosomal mCA presence. The y-axis shows –log10(*P*). Red points indicate associations significant after Bonferroni correction (*P* < 3.4 × 10^-5^). (B) Forest plot showing the association between autosomal mCAs (overall and high-risk CLL-associated) and protein levels. Beta estimates represent the change in standardized protein levels (SD units) per 10% increase in clonal fraction. All analyses were adjusted for age, age squared, sex, genetic ancestry, and smoking status.

### Risk of incident blood cell abnormalities of autosomal mosaic chromosomal alterations

Prior work has found that individuals with mCAs are more likely to have abnormal blood counts; however, the future risk of developing blood count abnormalities conferred by an mCA that is necessary for clinical counseling of patients with mCAs and for the design of clinical trials, is presently unknown. To quantify the risk of incident blood count abnormalities, we constructed a matched case-control cohort of 3,223 people with unique longitudinal blood counts data in UKB and AoU (**Supplementary Table 15**). The median time at risk was 1.6 years in those with aut-mCAs and 2.8 years in those without mCA. Incident persistent cytoses (defined by repeated abnormalities in blood counts ≥120 days apart) were two-fold more likely in those with aut-mCAs than those without. Cytoses occurred in 99/837 (11.8%) participants with aut-mCAs and 143/2,386 (5.9%) people without aut-mCAs. The major cause of cytosis was lymphocytosis, with 85 (10.2%) in mCA cases and 113 (4.7%) in controls. Those with aut-mCAs had significantly increased risk of incident cytosis versus matched controls (Hazard Ratio [HR]: 2.3, 95% CI 1.8-2.9, *P* = 1×10^-11^) (**Figure 4A**) (**Supplementary Table 16**). In contrast, the risk of incident cytopenia was not significantly different between those with and without aut-mCAs (HR: 1.1, 95% CI 0.96-1.30, *P* = 0.16) (**Supplementary Table 17**).

**Figure 4.**
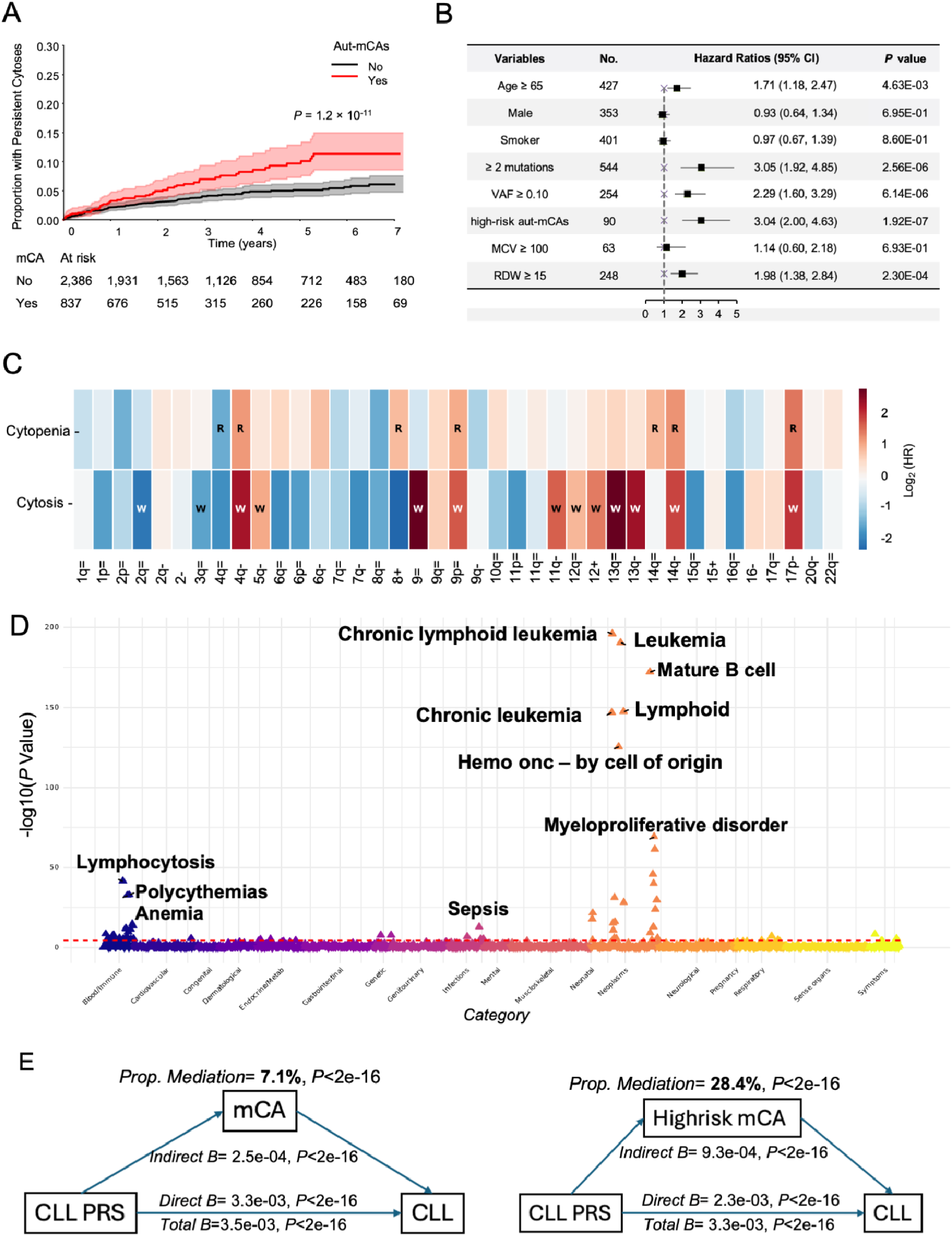
Clinical implications of autosomal mCAs. A) Cumulative incidence of cytosis comparing aut-mCA carriers (red) to matched controls (gray), accounting for competing risk of hematologic malignancy. Shaded areas represent 95% confidence intervals. The Fine- Gray model was used to estimate the cumulative incidence of cytosis. For those who did not develop hematologic malignancy, or persistent blood count abnormalities, the last follow-up date was defined by the latest time for CBC measurements. B) Forest plot showing hazard ratios (HR) for cytosis risk factors in aut-mCA carriers. Key risk factors include multiple mutations (HR=3.05, *P*=2.56×10^-6^) and high cell fraction (HR=2.65, *P*=5.10×10^-7^). The reference group for each association was defined as the other group of the trait. VAF, variant allele fraction; MCV, mean corpuscular volume; RDW, red cell distribution width. C) Heatmap showing associations between specific aut-mCA types and blood count abnormalities. Color intensity indicates effect size (log_2_HR). “W” indicates white blood cell, “R” red blood cell associations. D) Manhattan-style plot showing phenome-wide associations with aut-mCAs across disease categories after meta-analysis. Each point represents a phenotype, with −log_10_(P) values on y-axis. Upward/downward triangles indicate risk/protective associations. E) Mediation analysis showing aut-mCAs mediate 7.1% of CLL-PRS effect on CLL risk and F) high-risk CLL- associated aut-mCAs mediate 28.4% of CLL-PRS effect on CLL risk. The mediation model was used after adjusting for age, age squared, sex, current smoking status and cohort.

Risk factors for incident persistent cytoses included multiple aut-mCAs, CF ≥ 0.10, high-risk CLL-associated aut-mCAs, and red cell distribution width ≥ 15% (**Figure 4B)**. The presence of three or more of the aforementioned risk factors greatly increased risk of cytosis. In those in UKB with aut-mCAs and three or more risk factors, the cumulative incidence of cytosis over 2 years was 50% (37% - 66%) compared to 3% (1% - 8%) for those without any high risk features. Similarly, those with aut-mCAs and three or more risk factors in AoU, had a cumulative incidence of cytosis of 47% (44% - 51%) over 2 years (**Supplementary Table 18**).

Furthermore, specific aut-mCAs predisposed individuals to incident cytosis and/or cytopenias (**Figure 4C, Supplementary Table 19**). Persistent anemia and leukocytosis were the most common blood abnormalities. Consistent with their established roles in lymphoid malignancy, several high-risk CLL-associated aut-mCAs, including chr11q–, chr12+, chr13q–, and chr13q=, were associated with an increased risk of incident leukocytosis.^22^ Notably, our analyses extend prior observations by demonstrating subtype-specific patterns of hematologic dysregulation. For example, chr17q– increased risk of leukocytosis and anemia and chr6q– suggestively increased risk of cytopenia and decreased risk of cytosis. More broadly, we observed a clear divergence by lineage: myeloid-associated mCAs tended to increase risk of isolated persistent anemia without cytosis, and lymphoid associated mCAs tended to increase risk of isolated leukocytosis.

### Phenotypic consequences of mosaic chromosomal alterations

We sought to systematically profile risk of disease conferred by aut-mCAs. Time-to-event phenome-wide association studies (PheWAS) across AoU, UKB, and BioVU identified significant associations between mCAs and 58 distinct Phecodes (*P* ≤ 0.05/3,311) spanning multiple disease categories (**Supplementary Table 20**). The majority of these significant associations were consistent across 1KG-EUR-like (**Supplementary Table 21**) and 1KG-AFR-like (**Supplementary Table 22**) individuals. Most significant associations clustered in neoplasm systems with the most pronounced associations being observed with leukemias, particularly CLL (**Figure 4D**). Among 22,839 individuals with mCAs and no CLL diagnosis at baseline, we observed 402 incident CLL events during follow-up over 15 years (HR: 20.3, 95% CI 16.7-24.7, *P* = 9×10^-197^) **(Extended Data Fig. 13)**. The risk of CLL development varied by aut-mCA characteristics, with high clonal fraction (≥10%) demonstrating substantially higher CLL risk (HR: 45.0, 95% CI 21.8-92.6, *P* = 6×10^-25^), compared to those with lower clonal fraction (HR: 9.0, 95% CI 7.0-11.5, *P* = 6×10^-67^). Moreover, individuals carrying high-risk, CLL-associated aut- mCAs showed a 100-fold increased CLL risk compared to those without an aut-mCA (95% CI 47.2-214.6, *P* = 8×10^-33^).

In PheWASes of specific aut-mCA types, we replicated previously established high-risk CLL-associated mCAs, including chr13q=, chr13q-, and chr12+.^22^ Notably, beyond these well-characterized associations with lymphoid malignancy, our analyses extend prior work by systematically identifying mCA subtype–specific associations with non-hematologic diseases. We observed that several mCA types were strongly associated with cardiovascular, cerebrovascular, and kidney diseases, as well as infectious outcomes, highlighting previously underappreciated pleiotropic effects of mCAs across organ systems. For example, chr11q= increased the risk of peripheral vascular disease by 4.5-fold (95% CI: 2.6-7.8, *P* = 5.7×10^−8^) and chr13q- was significantly associated with sepsis (HR: 5.4, 95% CI: 3.6-8.2, *P* = 2.9×10^−15^) (**Supplementary Table 23**). Additionally, chr9p=, which co-occurs with *JAK2* CHIP, was associated with a 2.6-fold increased risk of stroke (95% CI: 1.7–3.7, *P* = 1.8×10^−6^) and a five-fold increased risk of chronic kidney disease (95% CI: 3.1–8.5, *P* = 3.0×10^−10^). We further validated all statistically significant phenotypes using the Firth logistic regression model to ensure robustness. By integrating the results from both models, we defined chr9p=, chr11q= and chr20q= as high cardiovascular-risk mCAs; chr9p= as a high stroke-risk mCA; chr4q= and chr9p= as high kidney disease-risk mCAs; and chr3q=, chr6p=, chr9p=, chr11q= and chr13q- as high infection-risk mCAs.

### Autosomal mCAs partially mediate and enhance germline polygenic CLL risk

Since aut-mCAs are strongly associated with CLL and CLL has a high single-nucleotide-polymorphism-based heritability, we hypothesized that (1) aut-mCAs partially mediate the risk of CLL in those with high CLL polygenic risk and (2) risk of CLL is higher in those with aut-mCAs and a high CLL polygenic germline genetic background than in those with aut-mCAs without high germline genetic risk. We calculated polygenic risk of chronic lymphocytic leukemia (CLL-PRS) from Law et al^32^ in UKB, AoU, and BioVU (**Supplementary Table 24**). Individuals with the highest 20% of CLL-PRS risk had the highest prevalence in all age groups for aut- mCAs and high-risk CLL-associated aut-mCAs (**Extended Data Fig. 14**). Nine specific types of aut-mCAs were associated with CLL-PRS (**Supplementary Table 11**). Aut-mCAs mediated 7.1% (*P* < 2 × 10^-16^) of the total CLL-PRS effect between CLL-PRS and CLL (indirect effect β = 2.5×10^-4^, *P* < 2×10^-16^; direct effect β = 3.3×10^-3^, *P* < 2×10^-16^; total effect β = 3.5×10^-3^, *P* < 2×10^-16^) **(Figure 4E)**. In a separate mediation analysis restricted to individuals with either high-risk CLL-aut-mCAs or no mCA, high-risk CLL-associated aut-mCAs mediated 28.4% (*P*< 2×10^-16^) of the total CLL-PRS effect (indirect effect β = 9.3×10^-4^, *P* < 2×10^-16^; direct effect β = 2.3×10^-3^, *P* < 2×10^-16^; total effect β = 3.3×10^-3^, *P* < 2×10^-16^) **(Figure 4F)**. These findings suggest that aut- mCAs, particularly those highly associated with CLL, substantially mediate the relationship between germline genetic risk and subsequent CLL development.

We found that those with aut-mCAs and high (top 20%) germline polygenic risk of CLL had substantially higher risk of CLL than those with either aut-mCAs or high polygenic risk. The 157,395 participants without detectable aut-mCAs but with high polygenic risk were at modest increased risk of CLL (HR: 2.14, 95% CI = 1.43-3.20, *P* = 2×10^-4^). The 17,501 people with detectable aut-mCAs but not at high polygenic risk were at higher risk of incident CLL than those with only high polygenic risk (HR: 23.32, 95% CI = 15.61-34.84, *P* = 3×10^-53^). The 5,113 people with both detectable aut-mCAs and high polygenic risk had the highest risk of CLL (HR: 49.16, 95% CI = 24.07-100.41, *P* = 1×10^-26^). The joint risk analysis indicates synergy between high polygenic risk and detectable aut-mCAs on the hazard of developing CLL (**Supplementary Table 25**).

## Discussion

This large-scale analysis of aut-mCAs across 1.2 million diverse individuals from four large biobanks enabled us to profile the remarkable heterogeneity of aut-mCAs compared to other forms of clonal hematopoiesis such as loss of sex chromosomes or CHIP. Certain aut-mCAs exhibit demographic specificity by age and sex. The older male predominance of chr15+ and chr20q– is supported by mathematical modeling of clonal fitness showing higher clonal expansion rate of these aut-mCAs in males after age 40.^11^ Moreover, germline variants increase risk of specific aut-mCAs. Expectedly, variants on the chromosome affected by the aut-mCA modulate risk of mCA acquisition, particularly for copy-neutral loss of heterozygosities and losses. Our largest-ever GWAS of each mCA yielded 6 novel loci and the inclusion of diverse genetic ancestries enabled discovery of variants with higher minor allele frequency in 1KG-AFR-like populations (e.g., the cis association at *PIK3C3* with chr18q=). Moreover, longer genetically predicted telomere length increased the risk of certain aut-mCAs, which suggests that this subset of aut-mCAs may specifically impose a selection pressure for hematopoietic stem cells as they replicate. While high-risk aut-mCA types for hematologic malignancy and certain types for infection have been reported, we are the first to perform PheWAS on each type of aut-mCA.

Despite the heterogeneity of aut-mCAs, our findings provide shared insights into the biology and clinical implications of clonal hematopoiesis. Most fundamentally, we nominated candidate drivers for aut-mCAs by integrating recurrent minimal regions with known hematologic-malignancy genes, prioritizing proto-oncogenes in gains and tumor suppressors in losses consistent with their expected dosage direction; CN-LOH regions yielded candidates of either class. This pattern may apply to copy-number variants in non-cancerous tissues throughout the body. Moreover, the largest GWAS of aut-mCAs individually demonstrates that variants near genes involved in mitochondrial metabolism (*TMEM70*), autophagy and lineage commitment (*PIK3C3*), and cell-cycle regulation (*FBXO39*) alter risk of acquiring specific mCAs. Our GWAS of aut-mCAs in aggregate demonstrates that genetic variants associated with aut-mCA prevalence broadly modulate either clonal expansion or immunosurveillance across all aut-mCA types. Loh et al had previously identified the risk loci in *TERT* and *SP140* for detectable mCAs on any autosome.^14,15^ The leading variant in *TERT* rs7705526-C, which reduces risk of aut-mCAs, was also shown to be associated with reduced risk of CHIP and CLL.^10,32–35^ We discovered three novel associations in *MAD1L1*, *TCL1A*, and *ATP2A3* with autosomal mCA prevalence. *MAD1L1*, previously associated with risk of chr15+ events in Biobank Japan, encodes the MAD1 spindle assembly checkpoint protein; biallelic germline loss-of-function mutations induce a syndrome of aneuploidy with high tumor susceptibility across the body. ^16,36,37^ *TCL1A* has been previously implicated in clonal expansion of HSCs, for both CHIP and mCAs and this variant likely alters prevalence of all aut-mCAs by modifying clonal expansion.^21,38^ Finally, the variant near *ATP2A3* is accessible in HSCs.^36^ *ATP2A3* encodes an ATP-dependent calcium transporter which alters the differentiation of HSCs likely toward lymphoid differentiation; prior work has demonstrated the importance of intracellular calcium homeostasis in HSC differentiation and function.^39,40^

Along with the aforementioned biologic insights, our findings may inform future studies of CLL risk stratification and the biological pathways linking specific aut-mCAs to CLL development. Approximately 12% of individuals with aut-mCAs developed incident cytosis, predominantly lymphocytosis, within 2 years. Among those with multiple high-risk features—including multiple aut-mCAs, abnormal red cell morphology, expanded clonal fraction, and CLL-associated aut-mCAs—approximately half developed cytosis within 2 years of sequencing. Because autosomal mCAs frequently co-occur with monoclonal B-cell lymphocytosis (MBL)^22^, the established precursor to CLL, some individuals with an aut-mCA and normal blood counts may harbor MBL. Distinguishing whether the incident lymphocytosis we observe (∼12% over 2 years) is therefore consistent with progression of underlying MBL or evidence that the mCA itself induces lymphocytosis de novo will require lineage-resolved data at-scale. Another high-risk feature is germline polygenic risk, which synergistically interacts with somatic aut-mCAs, to increase CLL risk. The proteomic signature of expanded, high-risk CLL-associated aut-mCAs revealed several proteins whose levels were significantly elevated and which overlap with known drug targets for hematologic malignancy. For example, those with high-risk CLL-associated mCAs had increased levels of NPM1 and PARP1, which are known druggable targets for leukemia (i.e., menin and PARP1 inhibitors) but novel for lymphoid leukemia.^41–43^ High-risk CLL-associated aut-mCAs were also associated with higher levels of TNFRSF13B (TACI), which interacts with B-cell activating factor (BAFF) and a proliferation-inducing ligand (APRIL) to increase B cell proliferation and class switching. Given the role of TACI in promoting the expansion of clonal B cells, increased circulating TACI levels may reflect B-cell clonal biology shared across aut-mCAs, MBL, and CLL.^22^ Telitacicept and atacicept both block the interaction of TACI with BAFF and APRIL and are investigative therapies targeting B cell proliferation for systemic lupus erythematosus.^44^

Limitations of this study include the single time point assessment of aut-mCAs, the heterogenous methods for mCA detection across cohorts, and use of telomere length and CLL polygenic scores derived in individuals of European ancestry. In addition, the observed lower detection rate of aut-mCAs in 1KG-EAS-like individuals may be influenced by reduced haplotype phasing accuracy in East Asian populations and their underrepresentation in cohorts. Furthermore, analyses of mCA-CHIP co-occurrence may be influenced by differences in sequencing depth and clonal cell fraction across cohorts, which could affect the sensitivity for detecting both mCAs and CHIP mutations. Although we observed broadly consistent directions of effect across cohorts, limited sample sizes for certain gene–mCA pairs in individual datasets may have reduced statistical power and contributed to variability in statistical significance. Moreover, despite directional concordance in the MESA replication for the proteomic analysis, limited case numbers reduced statistical power to achieve replication at stringent thresholds, highlighting the need for larger proteomic datasets to confirm these associations. Finally, incident blood count abnormality analyses could only be performed in those with longitudinal blood counts, which may overestimate the rate of incident blood count abnormalities, as these blood counts were collected in the course of routine clinical care.

In conclusion, this study provides fundamental insights into somatic structural variants which occur in the blood and likely in tissues throughout the body. Moreover, we provide evidence for an approach to incorporate germline genetics, somatic mosaicism, and clinical phenotypes to prioritize high-risk individuals who may benefit from preventative interventions. This paradigm likely applies to premalignant lesions in tissues throughout the body.

## Data Availability

Individual-level sequence data, CHIP calls and polygenic scores have been deposited with UK Biobank and are freely available to approved researchers, as done with other genetic datasets to date. The genotypes and phenotypes of UKB and AoU participants are available by application to the UKB (https://www.ukbiobank.ac.uk/register-apply/) and AoU (https://allofus.nih.gov/), respectively. Instructions for access to UK Biobank data are available at https://www.ukbiobank.ac.uk/enable-your-research. The HapMap3 reference panel was downloaded from ftp://ftp.ncbi.nlm.nih. gov/hapmap/, GnomAD v3.1 VCFs were obtained from https://gnomad.broadinstitute.org/downloads, and VCFs for TOPMED Freeze 8 were obtained from dbGaP as described in https://topmed.nhlbi.nih.gov/topmed-whole-genome-sequencing-methods-freeze-8. Vanderbilt BioVU data are available through an application to the Vanderbilt Institute for Clinical and Translational Research (VICTR) BioVU Review Committee. More information is available at https://victr.vumc.org/biovu/.

## Acknowledgments

This work was supported by National Institutes of Health grant R01AG083736 (A.G.B., P.L.A., P.S.), National Institutes of Health grant R01HL117626, National Institutes of Health contract HHSN268201800002I, National Institutes of Health grant R01HL120393, National Institutes of Health grant U01HL120393, National Institutes of Health contract HHSN268201800001I, National Institutes of Health grant DP5OD029586 (A.G.B.) and National Institutes of Health grant T32GM007347 (Y.P.).

The MESA projects are conducted and supported by the National Heart, Lung, and Blood Institute (NHLBI) in collaboration with MESA investigators. Support for MESA is provided by contracts 75N92025D00022, 75N92020D00001, HHSN2682015000031/HSN26800004, N01-HC-95159, 75N92025D00026, 75N92020D00005, N01-HC-95160, 75N92020D00002, N01-HC-95161, 75N92025D00024, 75N92020D00003, N01-HC-95162, 75N92025D00027, 75N92020D00006, N01-HC-95163, 75N92025D00025, 75N92020D00004, N01-HC-95164, 75N92025D00028, 75N92020D00007, N01-HC-95165, N01-HC-95166, N01-HC-95167, N01-HC-95168, N01-HC-95169, UL1-TR-000040, UL1-TR-001079, UL1-TR-001420, UL1TR001881, DK063491, and R01HL105756. Funding for SHARe genotyping was provided by NHLBI Contract N02-HL- 64278. Genotyping was performed at Affymetrix (Santa Clara, California, USA) and the Broad Institute of Harvard and MIT (Boston, Massachusetts, USA) using the Affymetrix Genome-Wide Human SNP Array 6.0. The authors thank the MESA participants and the MESA investigators and staff for their valuable contributions. A full list of participating MESA investigators and institutions can be found at http://www.mesa-nhlbi.org.

## Author Contributions

These authors contributed equally: Kun Zhao and Yash Pershad. K.Z., Y.P., A.G.B., A.P.R., P.S. and P.L.A. conceived the study. X.M., K.Q., H.M.P., T.M. and R.W.C. contributed to statistical analysis design. N.K.K., J.B., A.K. and Y.X. contributed to study design. Y.P., C.V., K.v.B. and Y.L. performed mCA and CHIP detection. W.C. and L.X. performed PRS statistical analyses. B.T., L.R., S.S.R., J.I.R., R.E.G. and M.Y.M. contributed to MESA data acquisition and data analysis. K.Z., Y.P. and A.G.B. performed statistical analyses and wrote the manuscript. All authors reviewed and revised the manuscript. A.G.B. supervised the work.

## Extended Data Figures

**Extended Data Fig. 1.**
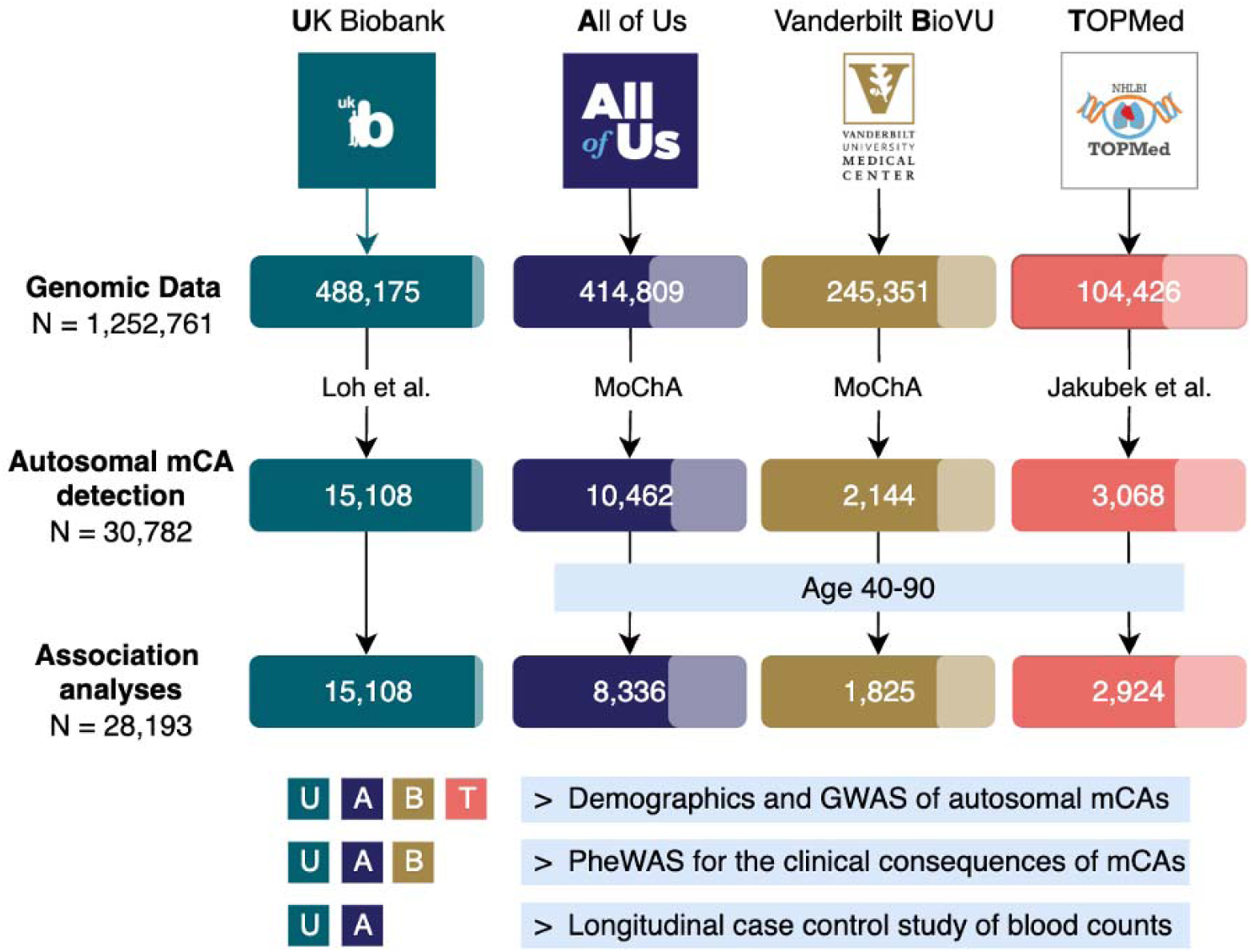
Study design and cohort selection. Flowchart showing the selection of participants from four large biobank cohorts: UK Biobank (N=488,175), NIH All of Us version 8 (N=414,809), Vanderbilt BioVU (N=245,351), and TOPMed (N=104,426). 30,782 individuals with autosomal mCAs were identified. Lighter sections indicate the proportion of non-European ancestry individuals. Bottom panels show which cohorts contributed to different analyses: demographics/GWAS (U,A,B,T), phenome-wide associations (U,A,B), and longitudinal blood count study (U,A).

**Extended Data Fig. 2.**
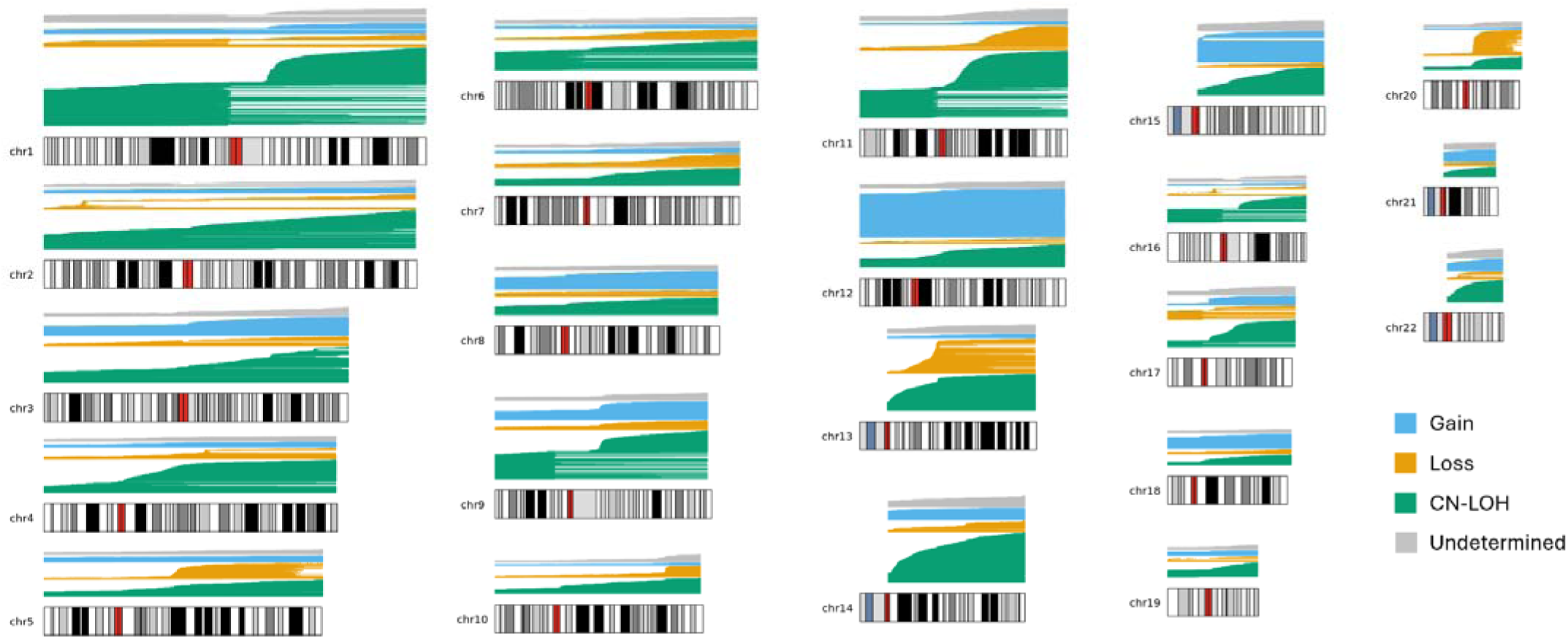
Genomic distribution of autosomal mosaic chromosomal alterations. Comprehensive visualization of aut-mCAs detected across autosomes in ∼1 million participants. Events are color-coded by type: gains (orange), copy-neutral loss of heterozygosity/CN-LOH (green), losses (blue), and undetermined copy number events (grey). Horizontal lines represent individual events, demonstrating the chromosomal locations and extent of detected alterations.

**Extended Data Fig. 3.**
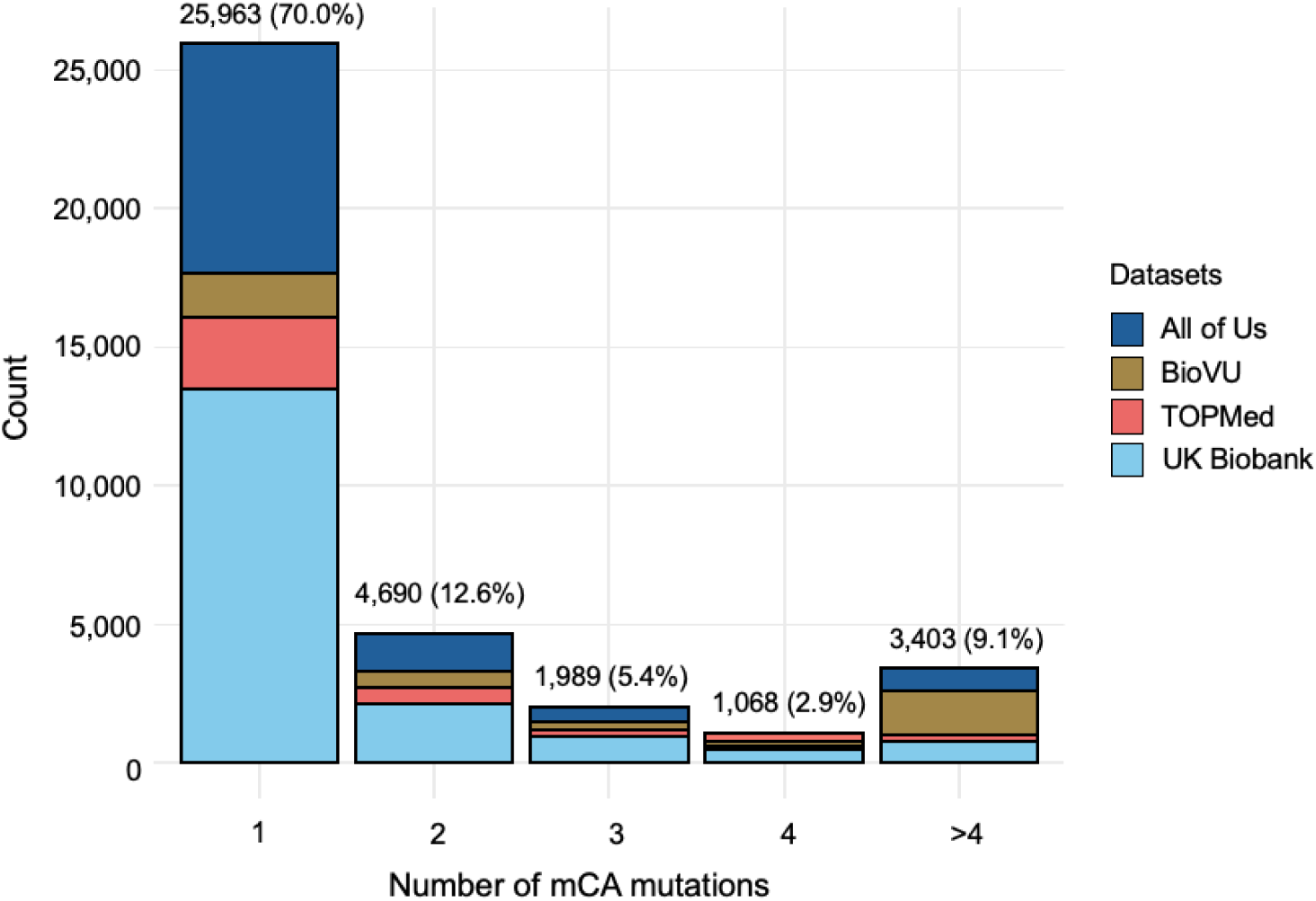
Distribution of the number of mCA mutations per carrier. Stacked bar plot showing the number of aut-mCA mutations per carrier across four cohorts. The majority (70.0%, N=25,963) carried a single mutation, while 12.6% (N=4,690) had two mutations, 5.4% (N=1,989) had three mutations, and 12.0% had four or more mutations. Data is stratified by cohort (UK Biobank, TOPMed, BioVU, and All of Us).

**Extended Data Fig. 4.**
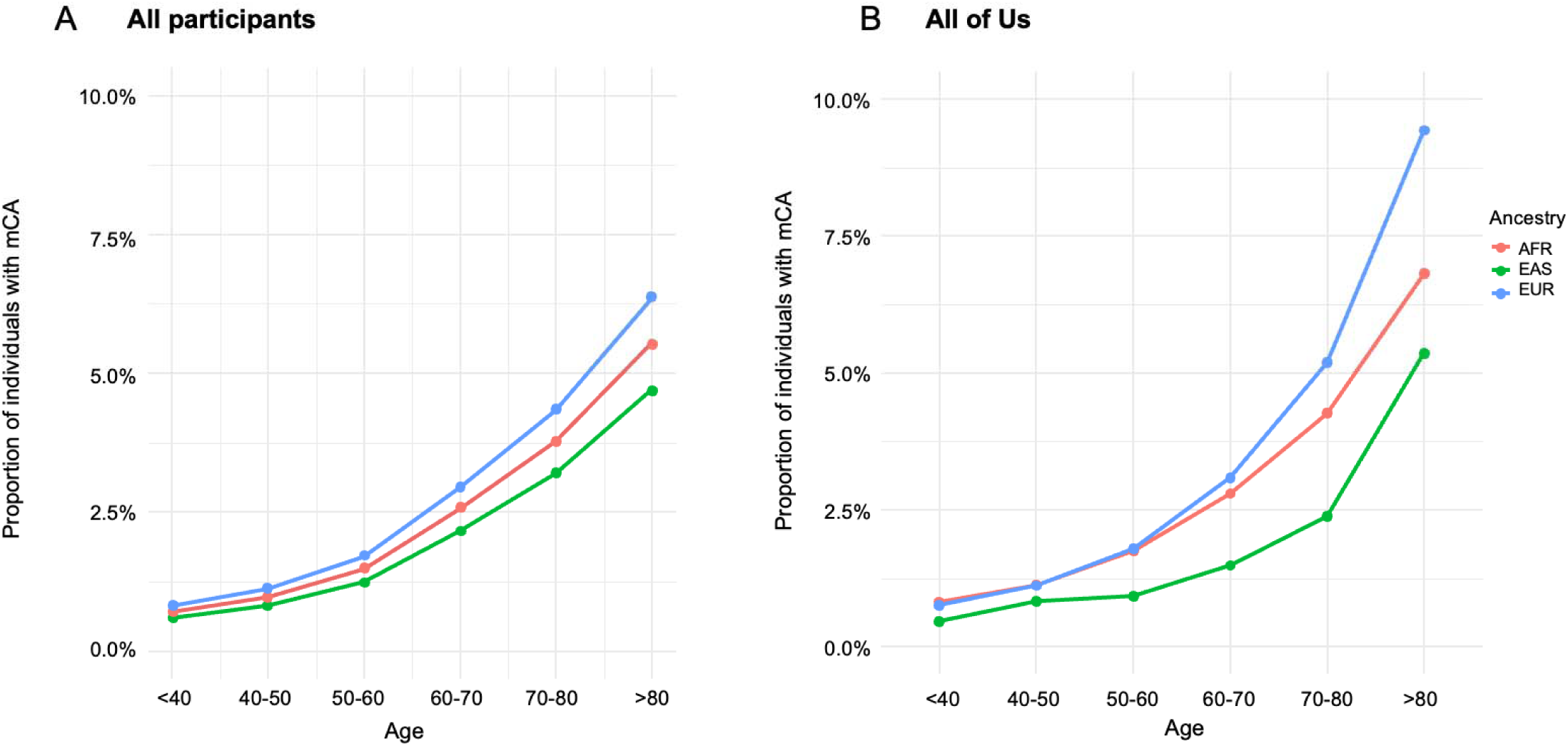
Age and ancestry-specific patterns of mCA detection rate. A) Overall aut-mCA detection rate across age groups stratified by genetic ancestry. B) Subset analysis in All of Us cohort showing similar patterns. Lines represent different ancestry groups: 1KG-AFR-like (red), 1KG-EAS- like (green), and 1KG-EUR-like (blue). Demonstrates increasing detection rate with age and ancestry-specific differences in accumulation rates.

**Extended Data Fig. 5.**
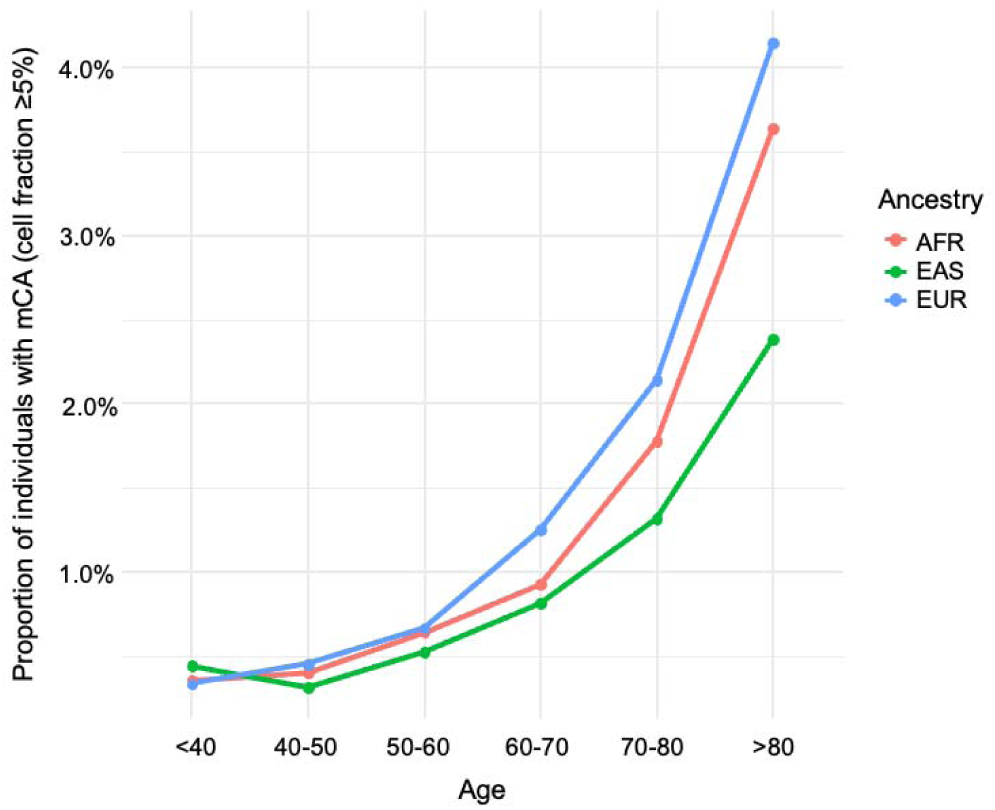
Age- and ancestry-specific patterns of mCA detection restricted to higher cell fraction events (CF >5%) Age- and ancestry-specific patterns of mCA detection restricted to higher cell fraction events (CF >5%). The detection rate of autosomal mCAs increases with age across all ancestry groups. Consistent differences between ancestry groups are observed, with 1KG-EUR-like and 1KG-AFR-like populations showing higher detection rates compared to 1KG-EAS-like populations. Lines represent different ancestry groups: 1KG-AFR-like (red), 1KG-EAS-like (green), and 1KG-EUR-like (blue).

**Extended Data Fig. 6.**
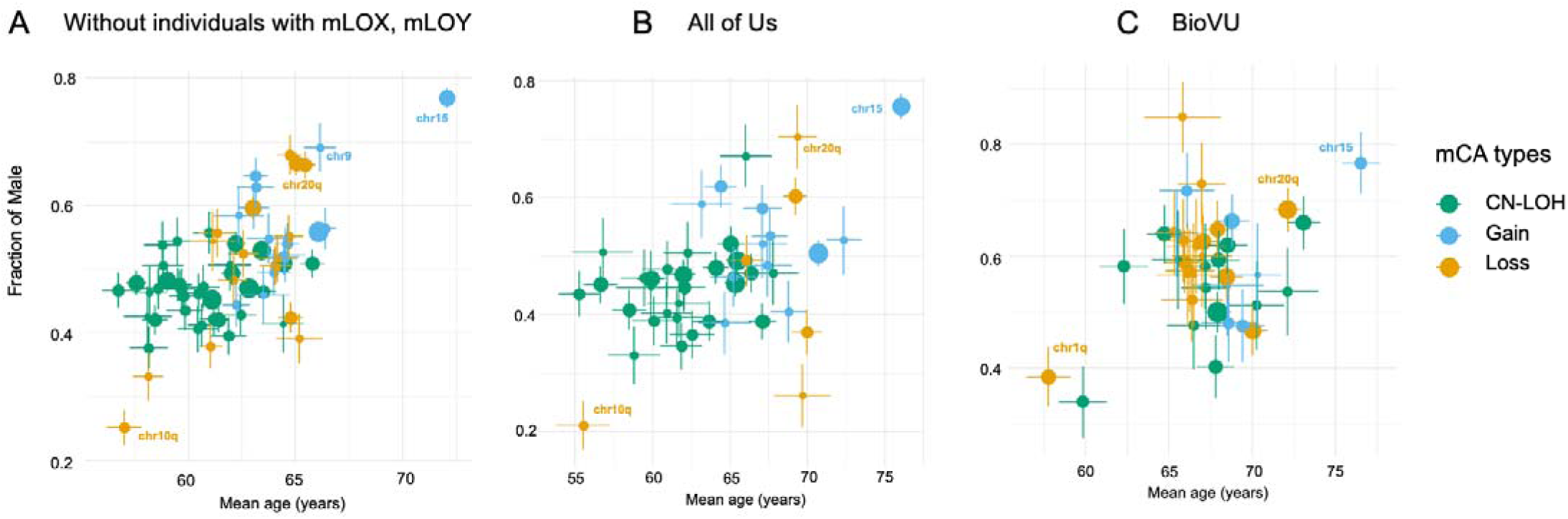
Sensitivity analysis of the sex and age distribution of mCA types. Scatter plot showing the relationship between mean age (x-axis) and proportion of males (y-axis) for different mCA types A) after excluding individuals with mosaic loss of chromosomes X or Y; B) in AoU cohort only; C) in BioVU cohort only. Point size indicates case count, colors denote mCA type (CN-LOH, gain, loss). Demonstrates that male predominance of chr15+ and chr20q- persists independently of sex chromosome mosaicism and is independently validated in both the AoU and BioVU cohorts.

**Extended Data Fig. 7.**
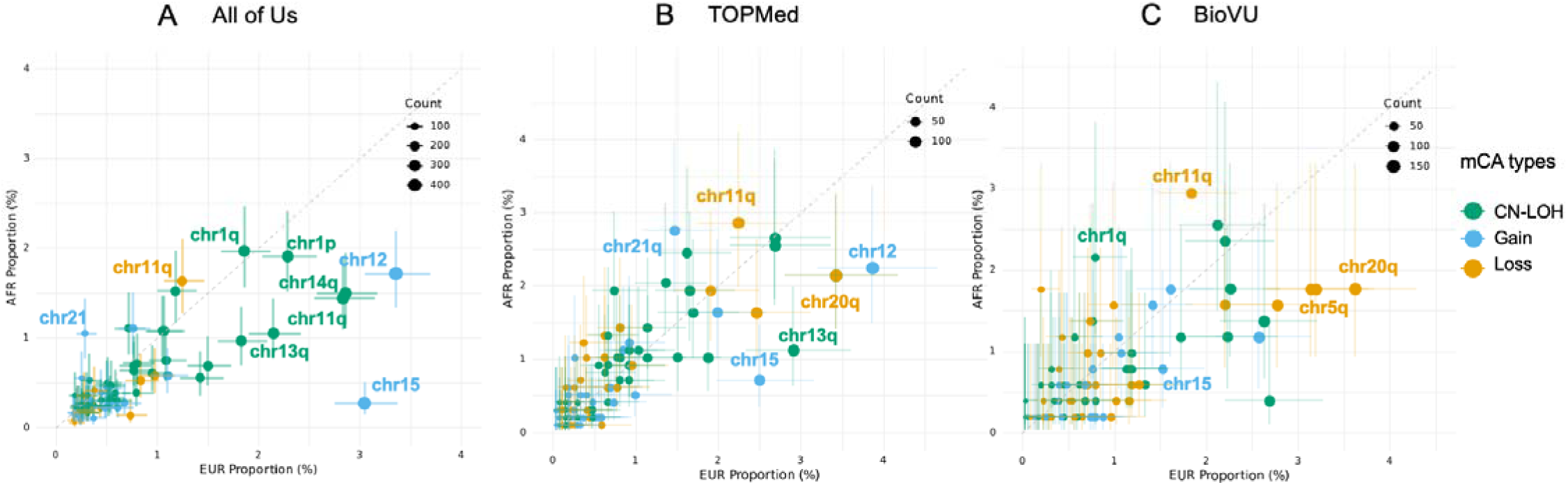
Distribution of ancestry for each aut-mCA types by cohort. Scatter plot showing the relationship between the proportion of 1KG-EUR-like (x-axis) and 1KG-AFR-like (y- axis) for different mCA types in A) AoU cohort ; B) TOPMed cohort; C) in BioVU cohort. Point size indicates case count, colors denote mCA type (CN-LOH, gain, loss).

**Extended Data Fig. 8.**
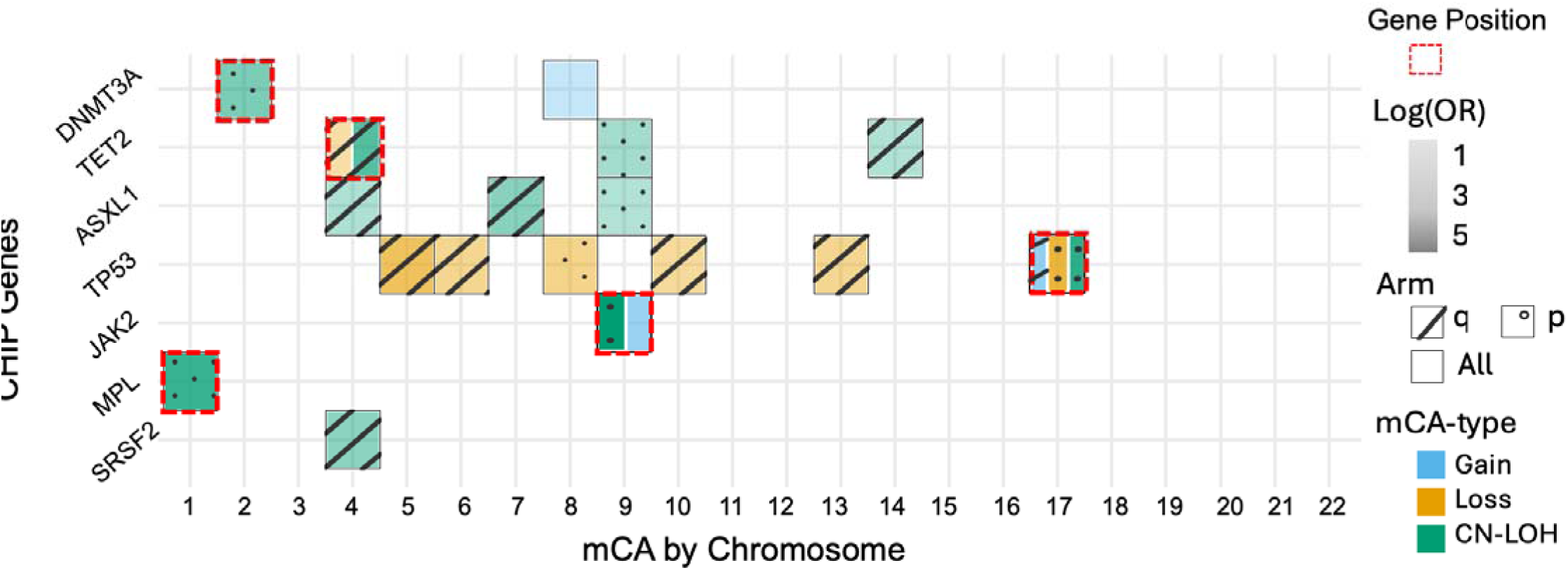
Co-occurrence patterns between CHIP mutations (VAF ≥5%) and aut-mCAs (CF ≥5%) across chromosomes. Color indicates aut-mCA type (CN-LOH, gain, loss). Firth logistic regression was performed and the model was adjusted for age, sex, and cohort to ensure robust results. Furthermore, co-occurrence pairs with fewer than 10 individuals were excluded, and a Bonferroni correction (*P* < 0.05/180) was applied to account for multiple testing.

**Extended Data Fig. 9.**
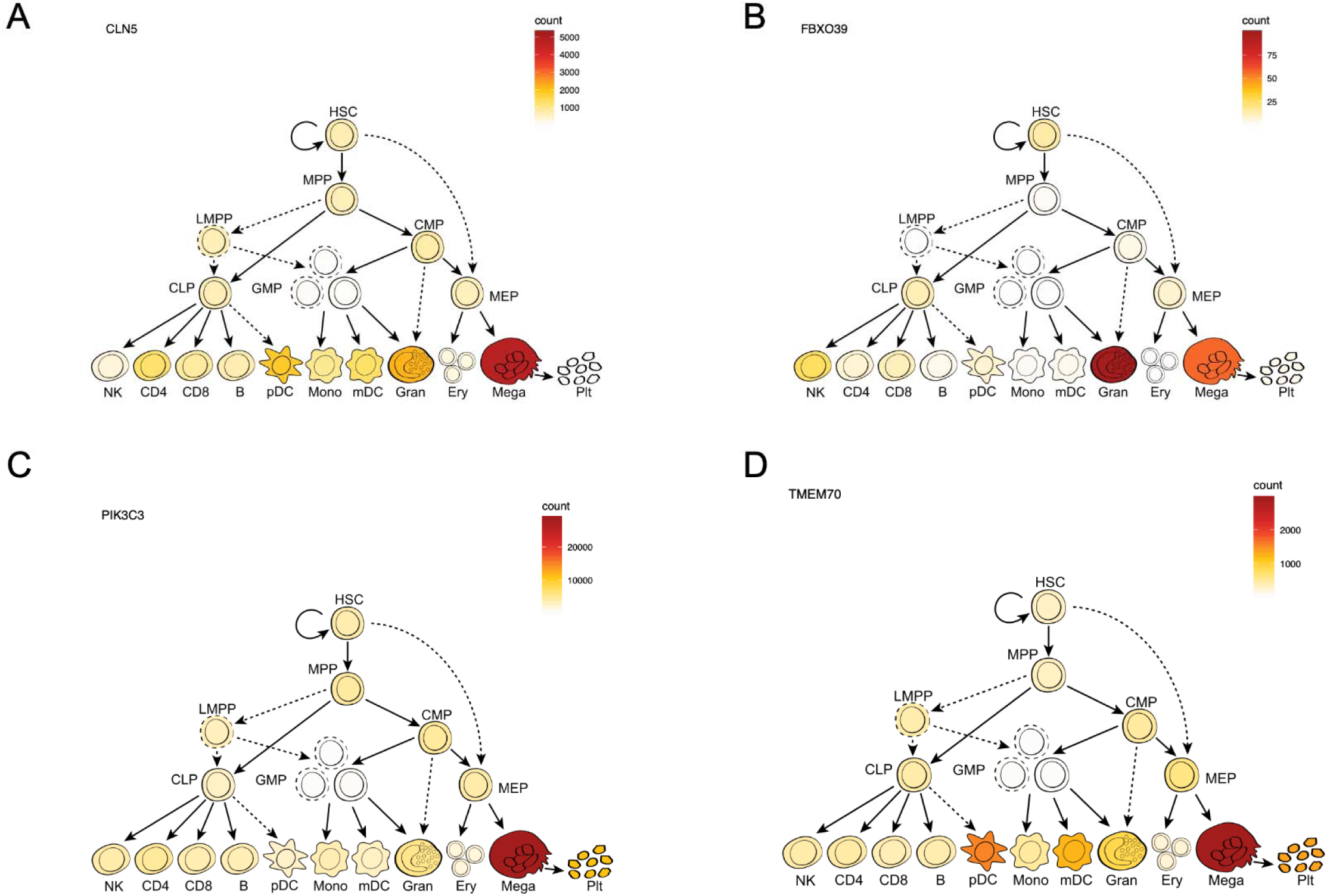
Hematopoietic expression patterns of genes implicated by novel germline associations with autosomal mCAs. Expression of CLN5 (**A**), FBXO39 (**B**), PIK3C3 (**C**), and TMEM70 (**D**) across hematopoietic stem and progenitor populations and differentiated blood-cell lineages. Color intensity represents gene-expression counts, with darker colors indicating higher expression. All four genes show expression in hematopoietic stem cells (HSCs), multipotent progenitors (MPPs), and common lymphoid progenitors (CLPs), with additional lineage- specific expression patterns. LMPP, lymphoid-primed multipotent progenitor; CMP, common myeloid progenitor; GMP, granulocyte– monocyte progenitor; MEP, megakaryocyte–erythroid progenitor; NK, natural killer cell; CD4, CD4 T cell; CD8, CD8 T cell; B, B cell; pDC, plasmacytoid dendritic cell; Mono, monocyte; mDC, myeloid dendritic cell; Gran, granulocyte; Ery, erythroid cell; Mega, megakaryocyte; Plt, platelet.

**Extended Data Fig. 10.**
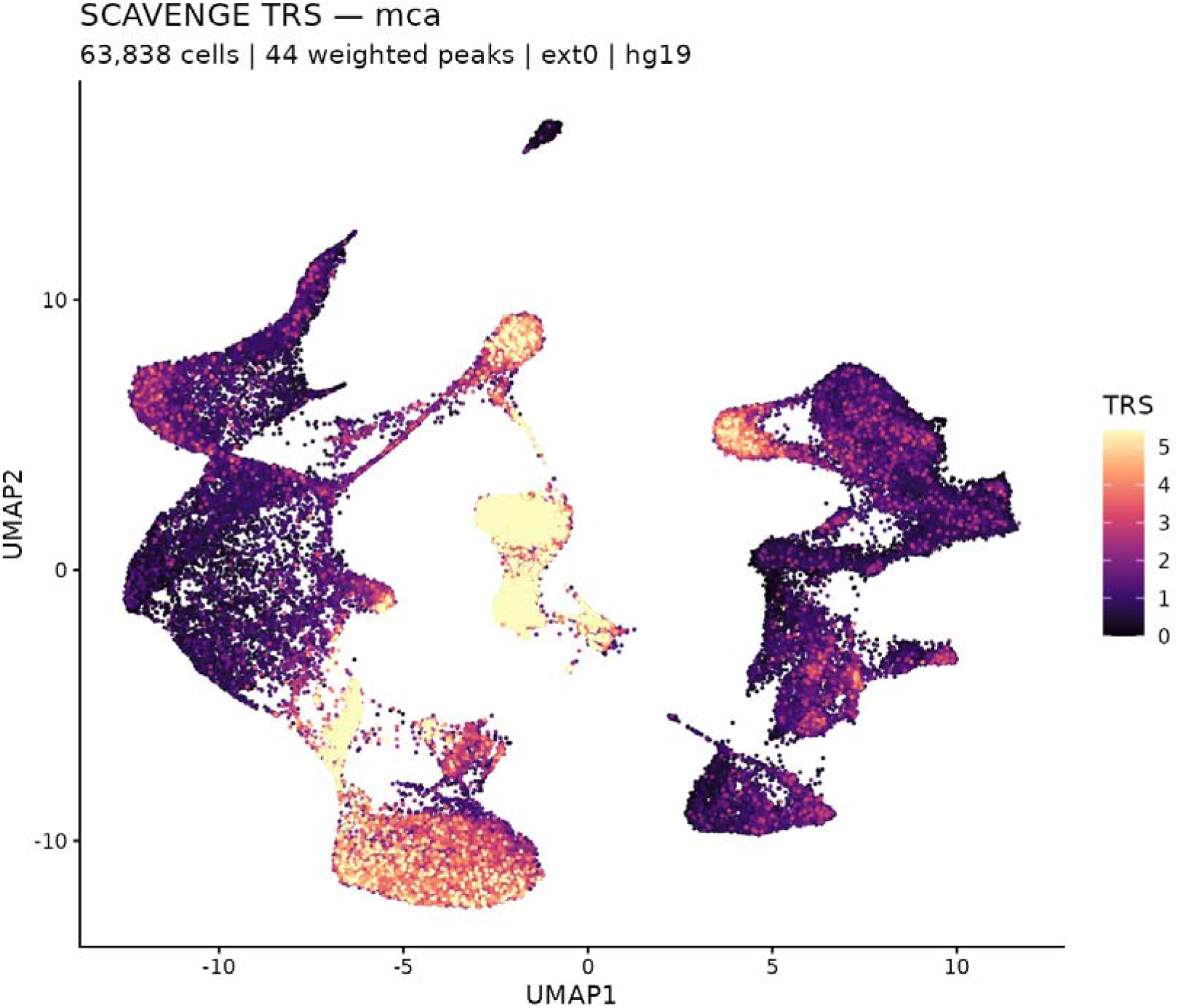
Single-cell chromatin enrichment of inherited genetic risk for autosomal mCAs. UMAP of 63,838 hematopoietic cells from Satpathy and Granja et al. single-cell chromatin accessibility reference, colored by SCAVENGE trait relevance score (TRS) for inherited aut-mCA risk. Higher TRS indicates greater enrichment of aut-mCA-associated genetic signal in accessible chromatin. The highest enrichment was shown for B cells (middle cluster).

**Extended Data Fig. 11.**
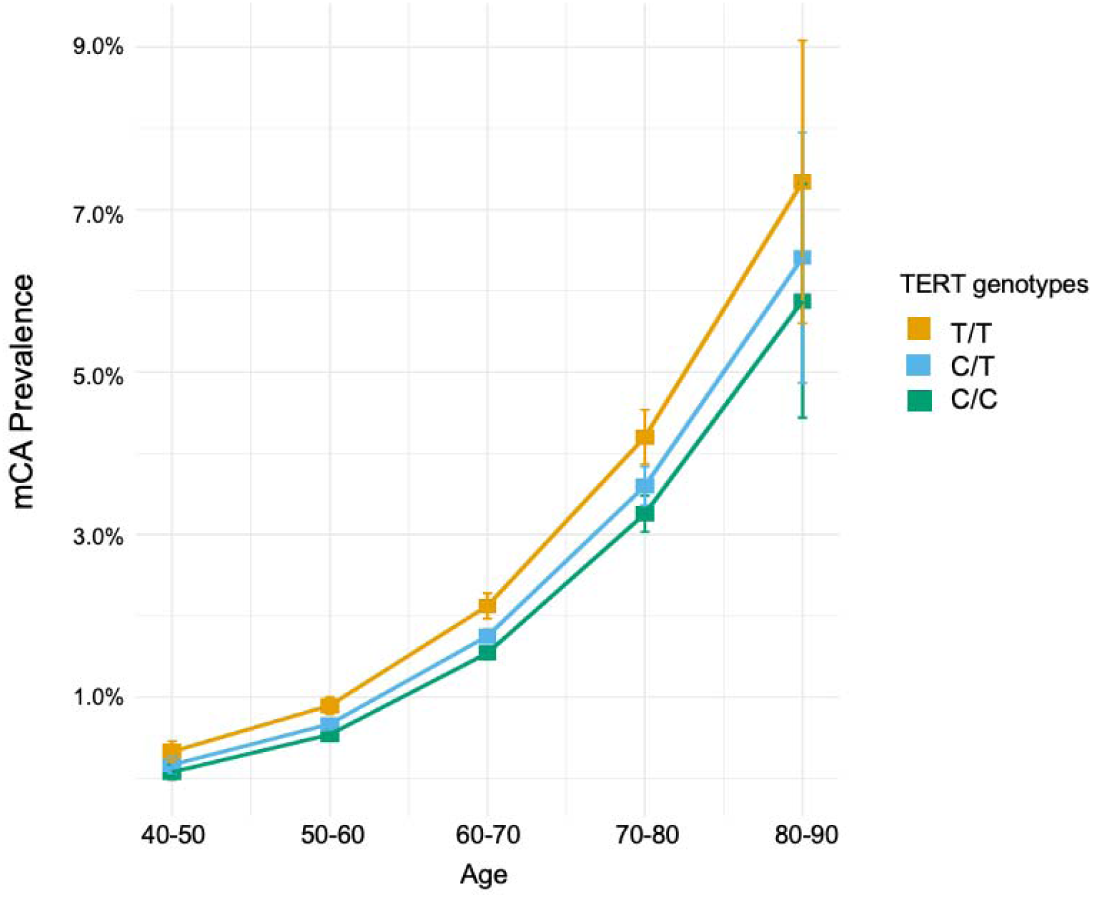
Relationship mCA prevalence and TERT genotype. The prevalence of aut-mCA stratified by TERT genotypes and age. Analysis restricted to European ancestry individuals from UKB, AoU, and BioVU. Shows consistently higher mCA prevalence in individuals with TERT T/T (orange) compared to C/T (blue) and C/C (green) groups.

**Extended Data Fig. 12.**
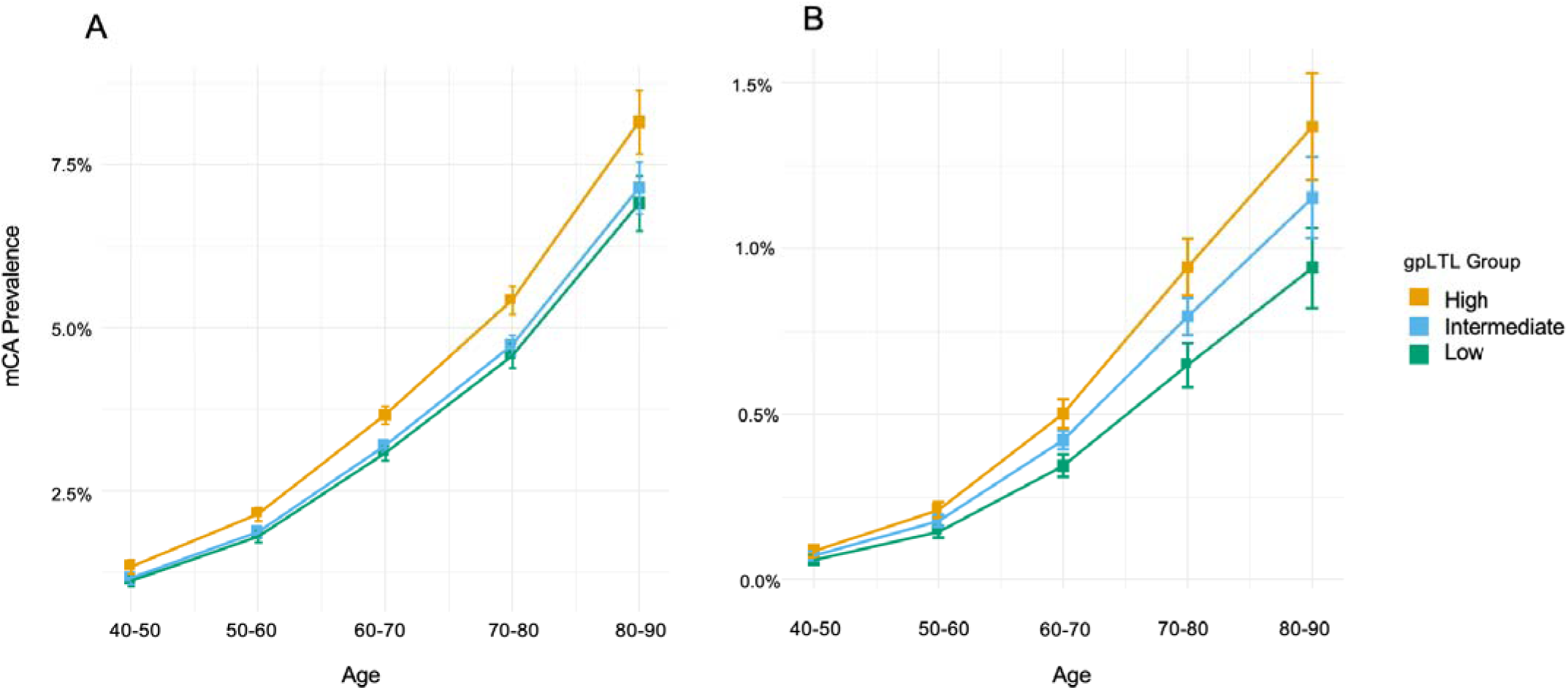
Relationship between genetically predicted leukocyte telomere length and mCA prevalence. A) Overall aut-mCA prevalence and B) high-risk CLL-associated aut-mCA prevalence stratified by genetically predicted leukocyte telomere length (gpLTL) groups and age. Analysis restricted to European ancestry individuals from UKB, AoU, and BioVU. Shows consistently higher mCA prevalence in individuals with high LTL-PRS (top 20%, orange) compared to intermediate (blue) and low (green) groups.

**Extended Data Fig. 13.**
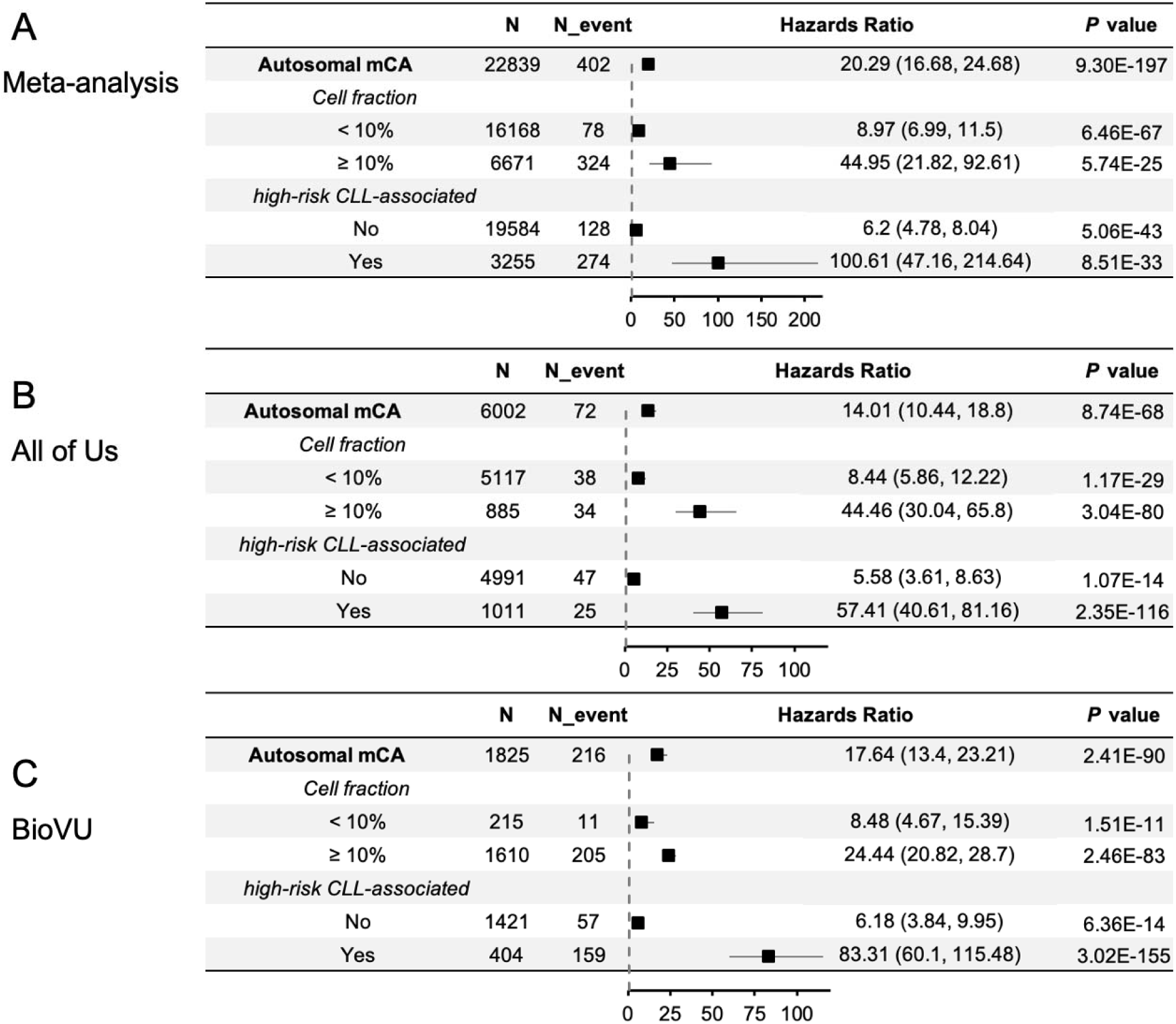
Impact of mCA characteristics on CLL risk. Forest plot showing hazard ratios for CLL incidence associated with different mCA features A) after meta- analysis across UKB, All of Us and BioVU; B) in All of Us cohort; C) in BioVU cohort. Demonstrates strong effects of cell fraction (≥10% HR=44.95 vs <10% HR=8.97) and CLL-risk classification (high-risk CLL- associated HR=100.61 vs non-high-risk CLL-associated HR=6.2) after meta-analysis. All analyses adjusted for age, sex, ancestry, and smoking status.

**Extended Data Fig. 14.**
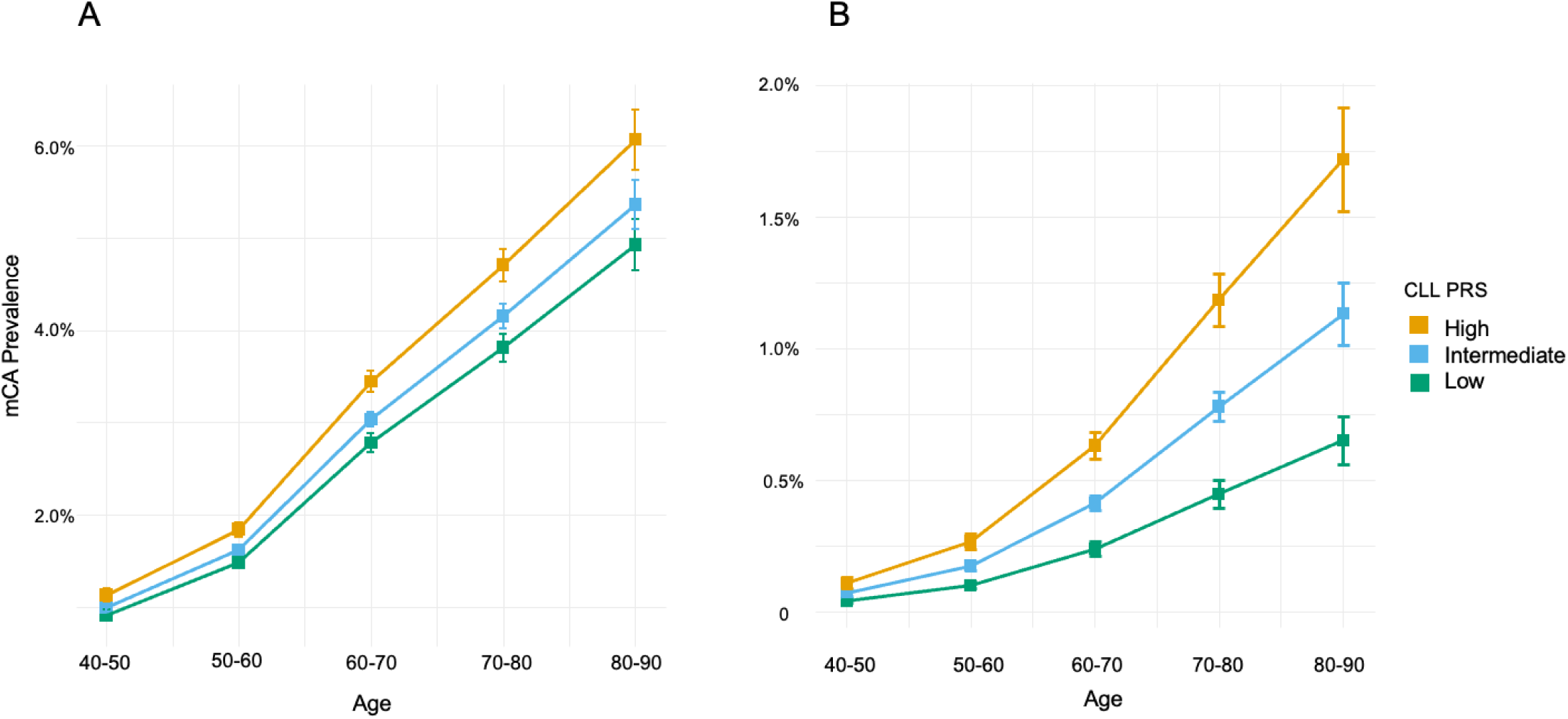
Relationship between CLL polygenic risk score and mCA prevalence. 1KG-EUR-like populations from UKB, AoU, and BioVU were included in the analysis. A) Age-specific aut-mCA prevalence stratified by CLL polygenic risk score (PRS) groups. B) Age-specific prevalence of high-risk CLL-associated aut-mCAs by CLL-PRS groups

