## Supplementary material for "Genetic drivers and clinical consequences of mosaic chromosomal alterations in 1 million individuals": Methods

#### Study cohort

This study utilized four datasets: the NIH All of Us (AoU) Research Program, UK Biobank (UKB), NHLBI Trans-Omics for Precision Medicine (TOPMed), and Vanderbilt’s BioVU. To ensure consistency across these datasets, participants aged 40–90 years were further analyzed (**Extended Data Fig.1**). The four cohorts are described as follows:

1) AoU aims to establish a longitudinal cohort of one million or more U.S. participants, with a particular focus on populations historically underrepresented in biomedical research. Detailed protocols for the AoU cohort and its genomic data have been described previously.^1,2^ In brief, eligible participants are U.S. residents or residents of U.S. territories aged 18 and older who can provide informed consent. The genotyping array data analyzed in this study were part of the AoU version 8 release and performed on the Illumina Global Diversity Array.

2) UKB is a population-based cohort of >500,000 UK adult residents recruited between 2006 and 2010 and followed prospectively via linkage to national health records.^3^ UKB included adults aged 40 to 70 years at blood draw with available genotyping array data (UK Biobank Axiom array). Follow-up in the UK Biobank occurred through March 2020 for inpatient diagnosis.

3) The NHLBI TOPMed program comprises data from over 51 studies, with whole-genome sequencing (WGS) conducted on all included samples, as previously described.^4^ The WGS data analyzed in this study were part of the TOPMed Freeze 8 data release.

4) BioVU is a Vanderbilt University Medical Center biobank with linked de-identified electronic health records (EHRs), spanning 2006 to 2021.^5^ Analyses were conducted using approximately 250,000 participants with available whole-genome sequencing (WGS) data. Sequencing libraries were prepared using the Illumina DNA PCR-Free Prep protocol and sequenced on Illumina NovaSeq 6000 instruments to a mean coverage of approximately 30×. The Vanderbilt University Medical Center’s Institutional Review Board oversees BioVU and approved this project.

#### Ancestry inference and analysis

Participants in AoU, BioVU, UKB, and TOPMed were assigned ancestry labels based on genetic clustering using principal components analysis within each cohort. Specifically, we computed principal components from a set of LD-pruned common autosomal SNPs using standard PCA procedures (FlashPCA2), and projected each participant’s genetic coordinates onto the 1000 Genomes (1KG) Phase 3 reference panel, which includes globally representative superpopulations (e.g., EUR, AFR, EAS). For consistency with other study cohorts and in accordance with the 2023 National Academies guidelines on population descriptors in genetics and genomics research^6^, we assigned individuals ancestry labels such as “1KG-EUR-like,” “1KG-AFR-like,” and “1KG-EAS-like” based on their genetic proximity to the 1KG reference clusters in principal component space. To ensure comparability across cohorts, we applied a uniform set of ancestry-informative, LD-pruned SNPs to all PCA pipelines where possible, and validated the resulting clusters against the known 1KG population structure.

For descriptive analyses (e.g., prevalence of mCAs subtype by ancestry), we performed cross-cohort analyses by pooling individuals with matched ancestry labels across cohorts (e.g., all 1KG-AFR-like individuals from UKB, AoU, TOPMed, and BioVU). For association analyses, we performed analyses within each cohort separately, adjusting for genetic ancestry via the first 5 principal components (PC1–PC5), along with age, age squared, sex, and smoking status. We then conducted meta-analyses across cohorts using either fixed-effects or random-effects models based on the degree of heterogeneity (I² statistic). The meta-analyzed results are reported in the main text.

#### Autosomal mosaic chromosomal alteration detection

We identified a total of 30,782 individuals with autosomal mosaic chromosomal alterations (aut-mCAs). In BioVU and TOPMed, mCA detection from whole-genome sequencing (WGS) data was performed using MoChA version 1.11, a haplotype phasing–based approach. MoChA leverages genotype information to estimate allelic balance and coverage at heterozygous loci, enabling detection of mCAs based on deviations in B-allele frequency (BAF) and read depth. Heterozygous markers were sourced from Taliun et al.^4^ and MoChA was run with the additional parameter ‘--LRR-weight 0.0 --bdev-LRR-BAF 6.0’, deactivating the LRR + BAF model to improve detection sensitivity. mCA detection in the UK Biobank was previously described.^7^ In brief, mCAs in blood DNA genotyping intensity data were detected using a statistical phasing-based approach validated in UKB. This method leverages long-range phase information to identify allelic imbalances, enhancing sensitivity for detecting large events at low cell fractions. Genotyping intensities were transformed to log2(R ratio) (LRR) and B-allele frequency (BAF) after affine-normalization and GC wave correction. In All of Us Research Program (version 8 genotyping data), detection of mCAs was performed starting from raw IDAT files generated using a single version of the Infinium Global Diversity Array, with MoChA version 1.11, following the same methods as applied in UKB and BioVU. We used the flag ‘-- adjust-BAF-LRR -1 --regress-BAF-LRR -1’ to avoid batch effects when processing array data. Across all cohorts, we focused exclusively on autosomal mCA events and excluded sex chromosome alterations (loss of X or Y). Detected mCAs were classified into copy-neutral loss of heterozygosity (CN-LOH), deletions, duplications, and events with undetermined copy number changes.

#### Quality control of mCA detection

mCA calls in UK Biobank and TOPMed were based on previously published pipelines. Given that UKB predominantly consists of individuals of European ancestry, we applied harmonized quality control procedures in the All of Us and BioVU cohorts, following approaches consistent with TOPMed to ensure comparability across datasets. Specifically, mCA calls were filtered to exclude low-confidence and likely artifactual events. These criteria included: (1) events spanning fewer than 2,000 informative heterozygous markers; (2) events with log-odds (LOD) scores <5; (3) events on chromosome X in individuals with uncertain inferred genetic sex; and (4) putative germline duplications identified based on thresholds of B-allele frequency (BAF) deviation and relative coverage. These filtering steps were designed to remove low-confidence calls and distinguish mosaic events from germline structural variation. At the sample level, standard quality control procedures were applied within each cohort, including evaluation of call rate, sex concordance, and relatedness. In addition, array-based datasets were processed using harmonized pipelines with BAF/LRR normalization and regression-based adjustment to reduce batch effects. Collectively, these QC procedures were applied consistently across cohorts to improve the robustness and comparability of mCA detection, particularly across ancestry groups.

#### Prioritization of candidate driver genes and enrichment analysis

To investigate the genetic drivers of aut-mCAs, we employed a two-stage approach: statistical enrichment of cancer gene classes and mapping of candidate genes within minimum shared altered regions (MSAR). First, to test if aut-mCA selection pressures follow known cancer genetics principles (as described by Davoli et al.^8^), we assessed the enrichment of tumor suppressor genes (TSGs) and oncogenes (OGs) within mCA regions. To test whether aut-mCA regions were enriched for cancer driver genes beyond chance, we performed a label-permutation test. We curated a list of experimentally validated hematologic malignancy genes from OncoKB and the MSK-IMPACT Heme panel.^9–11^ We then annotated these hematologic malignancy driver genes as tumor suppressors, proto-oncogenes, both, or neither using the OncoKB.^12,13^ For each aut-mCA type, the observed statistic was the number of recurrently lost regions containing ≥1 tumor suppressor and the number of recurrently gained regions containing ≥1 proto-oncogene; each region contributed at most one gene per class (presence/absence). We generated the null distribution by randomly permuting the tumor-suppressor/proto-oncogene annotations across all protein-coding genes 10,000 times while holding the genomic coordinates of both genes and mCA regions fixed, recomputing the statistic each time. Because gene positions are held constant, gene-dense loci (e.g., the LILR and MHC clusters) contribute identically under the null and do not generate spurious enrichment; the highly gene-dense, LD-complex MHC region (chr6:28,510,120–33,480,577, GRCh38) was excluded. Empirical P-values were (1 + #permutations with statistic ≥ observed) / (1 + 10,000). Enrichment was assessed using regions pooled across all four cohorts and, to test consistency, separately within UKB, AoU, BioVU, and TOPMed.

Second, to prioritize specific candidate drivers, we identified the minimum shared altered region (MSAR), the interval altered in a defined proportion of events of the same chromosome arm and type across all four cohorts. For each aut-mCA type we computed the region shared by 50%, 75%, 80%, and 90% of events and intersected each with UCSC hg38 gene coordinates. Restricting to curated hematologic malignancy genes, we selected, for each aut-mCA, the most stringent sharing threshold that retained ≥1 hematologic malignancy gene (thresholds in **Supplementary Table 5**). Candidate drivers were genes (1) within the selected MSAR and (2) biologically consistent with the copy-number change, proto-oncogenes for gains, tumor suppressors for losses; CN-LOH regions were not restricted by gene class. This prioritization defines candidate drivers by construction and is therefore descriptive rather than a statistical test of gene-class enrichment, which is assessed separately by the permutation test above.

#### Clonal hematopoiesis of indeterminate potential variant calls and co-occurrence analysis

CHIP and mCA events were detected from the same DNA source in each individual. Putative somatic SNPs and short indels were called with GATK Mutect2 (<https://software.broadinstitute.org/gatk>). Mutect2 first identifies candidate sites with evidence of variation and then performs local reassembly to refine variant calling. It employs an external reference, known as a “panel of normal samples,” to filter out recurrent sequencing artifacts and calls variants only at sites with evidence of somatic variation. The panel of normal samples comprised 100 randomly selected individuals under 40 years of age, with the absence of hotspot CHIP mutations confirmed prior to their inclusion. 74 canonical CHIP genes were screened for potential CHIP mutations using the Mutect2 somatic variant caller.^14^ Variants included in the preliminary dataset met the following criteria: presence in a pre-established list of candidate CHIP variants, total sequencing depth ≥ 20, alternate allele read depth count ≥ 5, and representation in both sequencing directions (i.e., F1R2 ≥ 1 and F2R1 ≥ 1). CHIP mutations were defined as those with a variant allele fraction (VAF) ≥ 0.02. The detail of CHIP calls in UKB, AoU, BioVU and TOPMed have been previously described.^15–17^ CHIP detection across all cohorts was derived from whole-genome sequencing data, using a consistent detection method and the same canonical CHIP driver genes list, which specifies candidate missense and indel variants for each gene, as well as a set of genes in which truncating and splice site variants may be considered.

We analyzed CHIP-mCA co-occurrence pairs using Firth logistic regression, a method that enables covariate adjustment while effectively handling small sample sizes. CHIP and mCA events were detected from the same DNA source in each individual. This analysis was restricted to individuals with at least one mCA and one CHIP mutation and evaluated whether certain mCA–CHIP gene pairs were enriched relative to all other possible pairs in this subgroup. The model was adjusted for age, sex, and cohort to ensure robust results. Furthermore, co-occurrence pairs with fewer than 10 individuals were excluded, and a Bonferroni correction (*P* < 0.05/180) was applied to account for multiple testing. Since MoChA can detect mCAs with CF as low as ~1-2%, while CHIP calling from WES typically requires a minimum VAF of ~5%, a sensitivity analysis was performed limited to co-occurring CHIP and mCA events where both had cell fractions/ VAF greater than 5% to ensure that both events are reliably detectable across platforms.

#### Genome-wide association studies

To identify germline variants associated with specific types of aut-mCAs, we conducted cis-GWAS analyses by restricting germline variants to those located on the same chromosomal arm as the corresponding mCA event. This design allowed us to evaluate local (cis) germline influences on somatic mosaic chromosomal alterations occurring within the same genomic region. Aut-mCA phenotypes on individual chromosome arms (p and q analyzed separately) were encoded as binary traits and included if at least 25 cases were available in each of the four cohorts (UKB, BioVU, AoU, and TOPMed). To control for population stratification, each cohort was stratified by genetic ancestry (1KG-EUR-like and 1KG-AFR-like, where sample size was sufficient) and analyzed separately.

In TOPMed, single-variant association testing was performed using SAIGE for variants with a minor allele frequency (MAF) >1% through the TOPMed Encore analysis server (<https://encore.sph.umich.edu>). In BioVU, AoU, and UKB, single-variant association testing was performed using REGENIE v3.3 with Firth correction for binary traits. A random subset of 500,000 variants with a minor allele count (MAC) >5,000 was used in step 1 of REGENIE. For step 2, variants with MAF <0.001 or a genotyping rate <0.9 were excluded. Participants without reported male or female sex at birth were also excluded. Across the cis-GWAS analyses, a total of 35,378,990 variant–phenotype tests were performed. Association models were adjusted for age, age², sex, smoking status, and 10 genetic principal components.

Ancestry- and cohort-specific association statistics were combined using fixed-effect, standard-error-weighted meta-analysis implemented in METAL (v2011-03-25). Test-statistic calibration was assessed using quantile–quantile (QQ) plots and genomic inflation factors (λGC). Because 81 chromosome arm-specific mCA phenotypes were tested, we applied a Bonferroni correction to the conventional genome-wide significance threshold (5 × 10^-8^), resulting in a study-wide significance threshold of *P* < 6 × 10^-10^.

Separately, we performed a genome-wide association study of overall aut-mCA prevalence, with the presence of any aut-mCA encoded as a binary phenotype. We used the same ancestry-stratified analytic framework, association software, covariates, and meta-analysis procedures described above. Because this analysis evaluated a single overall aut-mCA phenotype, variants were considered genome-wide significant at *P* < 5 × 10^-8^.

#### SCAVENGE

We applied SCAVENGE (v1.0.2, <https://github.com/sankaranlab/SCAVENGE>) to identify trait-relevant cell states for aut-mCA susceptibility. Fine-mapped variants from the aggregate aut-mCA meta-analysis (aBF, ω = 0.04, ±250 kb windows, variants tested in ≥2 cohorts) were used to construct a trait BED weighted by posterior inclusion probability. Variants were intersected with peaks from the Granja and Satpathy et al.^18^ human hematopoiesis scATAC-seq reference (hg19), enrichment was computed by gchromVAR, and seed cells were propagated across the cell–cell similarity network to yield a per-cell trait-relevance score (TRS). Per-cell significance was assessed against permuted background.

#### Locus definition and fine-mapping

Independent loci were defined by greedy distance-based pruning of the meta-analysis: the minimum-P variant was selected and all variants within ± 1 Mb on that chromosome removed, iterating until no variant below the significance threshold remained. This collapses multi-signal regions (e.g. *ATM*, *MPL*, *FRA10B*) to a single locus. No LD reference panel was used, because no single panel is appropriate across the contributing ancestry strata; for the same reason, fine-mapping methods requiring in-sample LD were not applied. For each locus we fine-mapped all variants within ± 1 Mb using the approximate Bayes factor method (prior variance ω = 0.04) to compute posterior inclusion probabilities and 95%/99% credible sets, yielding a single fine-mapped index variant per locus. Sensitivity to ω was assessed by repeating fine-mapping across a range of prior variances (e.g., 0.16).

#### Allelic shift analysis for cis-association validation

To test whether the chromosome carrying a cis risk allele is preferentially retained/amplified in the mCA clone, we used phased genotypes from All of Us v8 and TOPMed. For each lead cis variant, we restricted to mCA carriers heterozygous at the variant and used phase to determine, per individual, whether the mCA amplified/retained the effect or the alternate allele. We counted carriers in which the effect allele was over- versus under-represented (this reflects the relative phase of the variant and the mCA and is distinct from the raw VAF) and tested departure from the null of 0.5 with a two-sided binomial test, following the reference from Jakubek et al.^20^ Over/under counts and binomial P-values are reported per cohort and in meta-analysis (**Supplementary Table 6**), and calibration was confirmed with a QQ plot of allelic-shift P-values.

#### Proteomics association study

A total of 1,465 proteins were tested in 52,705 participants in the UK biobank. Proteomics was measured by Olink (Olink Proteomics; Uppsala, Sweden) using a proximity-extension immunoassay-based method, including proteins from cardiovascular, inflammation, cardiometabolic, neurology, oncology, and other panels. The TOPMed MESA (Multi-Ethnic Study of Atherosclerosis) cohort was used as a validation cohort using OLINK proteomic data (184 cases and 3,966 controls). MESA is a National Heart, Lung, and Blood Institute–sponsored prospective study aimed at studying the prevalence, progression, determinants, and prognostic significance of subclinical cardiovascular disease in a sex-balanced, multiethnic, community-dwelling U.S. cohort. Linear regression models were fitted with aut-mCAs (lymphoid and high-risk CLL-associated as described in **Supplementary Table 9**) or clonal fraction of aut-mCAs as exposures, the level of proteins as outcomes, and various covariates, including age at blood draw (continuous), age squared (continuous), genetic sex (categorical), current smoking status (categorical), and principal components (continuous). Linear regression models were performed using the R function ‘glm’. The Bonferroni threshold of P value was defined by 0.05/1,465 = 3.41 × 10^-5^. To evaluate the consistency of associations in MESA, we performed a sign test to assess whether proteins nominally associated in UKB demonstrated directional concordance and statistical significance in the validation cohort.

#### Genetically predicted LTL and Polygenic risk score of CLL

The genetically predicted LTL (gLTL) and polygenic risk scores for CLL were calculated for the 1KG-EUR-like population in the AoU, UKB and BioVU. To compute the scores, we first identified representative SNPs from each of susceptibility loci based on the most recent and comprehensive GWAS of CLL and LTL.^21,22^ A total of 41 SNPs and 131 SNPs were used for generating European-specific CLL-PRS and gLTL, respectively **(Supplementary Table 8, 24)**. The PRS/ gLTL were calculated by summing all variants with the following formula:

$$CLL PRS/gLTL = \sum_{i = 1}^{n} \beta i \cdot SNPi$$

Where *βi* represents the estimated weight (i.e., the natural logarithm of the odds ratio [OR]) of the i-th SNP, derived from the reference datasets, and *SNPi* is the genotype dose of each risk allele for that SNP.^23^ CLL PRS/ gLTL values were categorized into low (<20%), intermediate (20% ≤ PRS/ gLTL <80%), or high (≥80%) genetic risk groups. To assess the association between CLL PRS/ gLTL and aut-mCA incidence, Firth logistic regression models were employed to estimate ORs and 95% CIs, adjusting for age at blood draw, age squared, genetic sex, smoking status and cohort.

#### Incident blood count abnormality analysis

We conducted a case-control study of participants from AoU and UKB. Individuals were eligible for the study if they had sequencing/genotyping for mCA detection and longitudinal complete blood count (CBC) data, without evidence of cytopenia or cytosis, acute myeloid leukemia (AML), myelodysplastic syndrome (MDS), myelofibrosis, or CLL prior to sequencing. Longitudinal CBC was defined as at least three CBC measurements, including one within a year of sequencing and two on or after the date of sequencing. The final CBC measurement had to occur at least 120 days after sequencing or the first CBC measurement, whichever came later. CBC measurements occurring greater than one year before sequencing were not included in the analysis. Participants with autosomal mCAs were matched 1:3 with controls on age, sex, and smoking status. The follow-up period commenced at the date of sequencing and terminated at the earliest occurrence of myelofibrosis, MDS, AML, CLL, or death.

Cytosis were defined by using a modified version of World Health Organization criteria (anemia: hemoglobin > 16.5 g/dL (females) or 18.5 g/dL (males); thrombocytopenia: platelets > 450,000 cells/mL; and leukopenia: white blood cell count < 11,000 cells/mL). Cytopenias were defined by using a modified version of World Health Organization criteria 8 (anemia: hemoglobin < 12.0 g/dL (females) or 13.0 g/dL (males); thrombocytopenia: platelets < 150,000 cells/mL; and leukopenia: white blood cell count < 3,700 cells/mL). Cytosis and cytopenias were only deemed to be *persistent* if there were two consecutive observations of a cytosis in a single lineage at least 120 days apart without an intervening normal measurement. The date of cytosis was the first occurrence of the cytosis that persisted for at least 120 days.

The primary outcome of interest was incident cytosis any time after study enrollment. Date of birth and death, sex, race, laboratory values, smoking status, and ICD-9/ICD-10 codes were extracted. All laboratory measurement variables were harmonized to a common unit of measure and screened for outlier values. Cumulative incidence of cytosis were estimated by using the Fine-Gray model due to the potential competing risk of hematologic malignancy. For those who did not develop hematologic malignancy, or persistent blood count abnormalities, the last follow-up date was defined by the latest time for CBC measurements. Survival time was calculated by the time difference between the last follow-up date and baseline date for the blood draw.

#### Phenome-wide association study

AoU, UKB, and BioVU cohorts, which have available outcome data, were included in the Phenome-wide association studies (PheWAS). We determined phenome-wide clinical outcomes based on PheCodes (https://phewascatalog.org/phewas/#phex) derived from International Classification of Diseases, Ninth or Tenth Revision (ICD-9/ICD-10) codes curated from EHRs. For each subject, the presence of any ICD codes corresponding to a PheCode inclusion criterion classified the subject as a case, while the absence of these codes classified the subject as a control. In addition to overall aut-mCAs, we leveraged the large sample size of this study to investigate specific types of aut-mCAs. We selected mCA types with a sample size of ≥200 in at least one dataset, resulting in 18 types included in the analysis. For the PheWAS analyses in UKB, AoU, and BioVU, we applied Cox proportional hazards models (using the R package “survival”) to evaluate associations between mCAs (or specific mCA types) and the risk of various phenotypes, estimating hazard ratios (HRs) and 95% confidence intervals (CIs). For all disease association analyses, individuals with pre-existing diagnoses for the phenotype or hematologic malignancies prior to DNA collection were excluded. The models were adjusted for age at blood draw (continuous), age squared (continuous), genetic sex (categorical), current smoking status (categorical), and principal components (continuous). For the significant results from the PheWAS of specific mCA types, we performed a sanity check using Firth logistic regression. Only results that were significant in both models were reported. PheWAS analysis was performed by each dataset and combined through inverse variance-weighted, fix-effects or random-effects meta-analysis (R package “metafor”).

#### Mediation analysis

Mediation analyses (using the R package “mediation”) were conducted to evaluate whether mCA mediated the relationship between CLL-PRS and CLL. This approach estimates the total effect of CLL-PRS on CLL and decomposes it into the direct effect (the effect of CLL-PRS on CLL independent of aut-mCAs) and the indirect effect (the portion of the effect mediated through aut-mCAs). The mediation analysis was performed using a two-stage regression process: first, modeling the association between CLL-PRS and aut-mCAs (mediator model), and second, modeling the association between aut-mCAs and CLL while adjusting for CLL-PRS (outcome model). The average causal mediation effect (ACME) and average direct effect (ADE) were estimated, with statistical significance evaluated through bootstrapping. All models were adjusted for age, age squared, sex, smoking status, and cohort. Separate mediation analyses were conducted for (i) all autosomal mCAs and (ii) high-risk CLL-associated autosomal mCAs. In the high-risk analysis, individuals with low-risk mCAs were excluded, such that the mediator compared high-risk mCA carriers to individuals without mCAs. As a result, these analyses were conducted in partially overlapping but non-identical analytic samples and used different mediator definitions; therefore, the estimated proportions mediated should not be interpreted as a strict subset decomposition. The mediation analysis relies upon the assumption that there is not a confounding variable that increases both risk of aut-mCA and CLL. Given that mCAs are implicated in clonal expansion and genomic instability—key processes in CLL pathogenesis—it is plausible that aut-mCAs act as intermediaries linking genetic predisposition (via CLL-PRS) to the development of CLL.

### **Code availability**

The code is publicly available and can be found at https://github.com/bicklab/mca-1m. The REGENIE software is available at https://github.com/rgcgithub/regenie. A standalone software implementation (MoChA) of the algorithm used to call mCAs is available at https://github.com/freeseek/mocha.

### **Data availability**

Individual-level sequence data, CHIP calls and polygenic scores have been deposited with UK Biobank and are freely available to approved researchers, as done with other genetic datasets to date. The genotypes and phenotypes of UKB and AoU participants are available by application to the UKB (https://www.ukbiobank.ac.uk/register-apply/) and AoU (https://allofus.nih.gov/), respectively. Instructions for access to UK Biobank data are available at https://www.ukbiobank.ac.uk/enable-your-research. The HapMap3 reference panel was downloaded from ftp://ftp.ncbi.nlm.nih. gov/hapmap/, GnomAD v3.1 VCFs were obtained from https://gnomad.broadinstitute.org/downloads, and VCFs for TOPMED Freeze 8 were obtained from dbGaP as described in <https://topmed.nhlbi.nih.gov/topmed-whole-genome-sequencing-methods-freeze-8>. Vanderbilt BioVU data are available through an application to the Vanderbilt Institute for Clinical and Translational Research (VICTR) BioVU Review Committee. More information is available at <https://victr.vumc.org/biovu/>.
